# The emergence of SARS-CoV-2 lineages and associated antibody responses among asymptomatic individuals in a large university community

**DOI:** 10.1101/2023.01.30.23285195

**Authors:** Marlena R. Merling, Amanda Williams, Najmus Mahfooz, Marisa Ruane-Foster, Jacob Smith, Jeff Jahnes, Leona W. Ayers, Jose A. Bazan, Alison Norris, Abigail Norris Turner, Michael Oglesbee, Seth A. Faith, Mikkel B. Quam, Richard T. Robinson

## Abstract

SARS-CoV-2 (CoV2) infected, asymptomatic individuals are an important contributor to COVID transmission. CoV2-specific immunoglobulin (Ig)—as generated by the immune system following infection or vaccination—has helped limit CoV2 transmission from asymptomatic individuals to susceptible populations (e.g. elderly). Here, we describe the relationships between COVID incidence and CoV2 lineage, viral load, saliva Ig levels (CoV2-specific IgM, IgA and IgG) and inhibitory capacity in asymptomatic individuals between Jan 2021 and May 2022. These data were generated as part of a large university COVID monitoring program and demonstrate that COVID incidence among asymptomatic individuals occurred in waves which mirrored those in surrounding regions, with saliva CoV2 viral loads becoming progressively higher in our community until vaccine mandates were established. Among the unvaccinated, infection with each CoV2 lineage (pre-Omicron) resulted in saliva Spike-specific IgM, IgA and IgG responses, the latter increasing significantly post-infection and being more pronounced than N-specific IgG responses. Vaccination resulted in significantly higher Spike-specific IgG levels compared to unvaccinated infected individuals, and uninfected vaccinees’ saliva was more capable of inhibiting Spike function. Vaccinees with breakthrough Delta infections had Spike-specific IgG levels comparable to those of uninfected vaccinees; however, their ability to inhibit Spike binding was diminished. These data demonstrate that COVID vaccines achieved hoped-for effects in our community, including the generation of mucosal antibodies that inhibit Spike and lower community viral loads, and suggest breakthrough Delta infections were not due to an absence of vaccine-elicited Ig, but instead limited Spike binding activity in the face of high community viral loads.

## INTRODUCTION

Coronaviruses are single-stranded RNA viruses that cause respiratory disease in a range of mammalian hosts. The Coronavirus Disease 2019 (COVID-19, or COVID) pandemic began in December 2019, after transmission of a novel coronavirus to an individual living in China. The sequence homology of this novel coronavirus to severe acute respiratory syndrome associated coronavirus (SARS-CoV) led to its being named SARS-CoV-2 (CoV2). CoV2 spreads via aerosol and respiratory droplets, causing either an asymptomatic infection or a flu-like illness that affects multiple organ systems and presents as fever, cough, dyspnea, malaise, delirium and death. International spread of CoV2 was rapid, and by February 2020 it had spread to nearly every country in the world (*1*). Now, 3 years after its emergence, CoV2 is estimated to have infected ∼630 million individuals and killed >6.5 million individuals worldwide (*2*). The United States has reported more deaths than any other country (*2*).

Viruses mutate to varying degrees depending on the nature of their genome and the proofreading activity (or lack thereof) of associated polymerases. CoV2 is no exception to this, and within a year of its emergence multiple lineage variants of concern (VOCs) appeared in numerous countries. B.1.1.7 (now called Alpha) and B.1.351 (now called Beta) were the first VOCs to be identified in September 2020 (Alpha, in United Kingdom) and October 2020 (Beta, in South Africa), and contained numerous missense mutations affecting the Spike protein (*3, 4*). The Spike protein is essential for CoV2 infection of target cells and contains a receptor-binding domain (RBD) which recognizes and binds the host receptor angiotensin-converting enzyme 2 (ACE2) (*5*). The Alpha and Beta lineage RBD mutations lead to tighter Spike:ACE2 structural interactions (*6*) and increased the transmissibility of CoV2 (*7, 8*). In January 2021, the P.1. (now called Gamma) lineage was reported in Brazil to contain even more missense mutations in more genes, including Spike (*9*). As with Alpha, the mutations inherent to the Gamma lineage increased its transmissibility (*9*). Two additional lineages emerged in March 2021 and November 2021, respectively, and in time would supplant all prior lineages in the speed with which they spread: the Delta lineage, which was first reported in India (*10*), and the Omicron lineage, reported in southern Africa (*11*). CoV2 continues to evolve, and deaths due to COVID continue to cause overall declines in life expectancy for many countries, including the United States (*12, 13*).

After previous coronavirus disease outbreaks, such as those caused by SARS-CoV and Middle East respiratory syndrome coronavirus (MERS-CoV), animal models and other experimental systems demonstrated that coronavirus-specific antibodies are generated soon after infection (*14, 15*), and can block viral entry by interfering with the Spike:ACE2 interaction (*16-21*). In the upper respiratory tract and oral cavity, antibodies are generated by B cells in mucosa-associated lymphoid tissue (MALT) and regional draining lymph nodes, typically within several days of antigen encounter, and comprise several isotypes (IgM, IgA and IgG) which differ in their secretion kinetics and effector mechanism. IgM is often the first isotype to appear following antigen exposure, and eliminates viruses by precipitating the membrane attack complex on virus-infected cells (i.e. the classical complement pathway). In the context of CoV2 infection, however, IgA dominates the early neutralizing antibody response at mucosal sites (*22*). IgA, a weak inducer of the complement pathway, protects mucosal sites by blocking and sterically hindering antigen interaction with the epithelial surface, trapping it in mucus which is eventually cleared via peristalsis. IgG is often the last isotype to appear following antigen exposure but is the most versatile in terms of effector mechanisms and durability, as the B cells which produce IgG can become plasma cells that reside in bone marrow and continuously secrete IgG for months to years.

The fact that coronavirus-specific Ig is secreted following natural infection, long-lived, and able to disrupt Spike:ACE2 interactions are the foundations on which multiple monitoring, therapeutic and vaccine strategies against CoV2 have been built. Prior to mass PCR testing, CoV2-reactive Ig in sera was the only biomarker for monitoring CoV2 prevalence at a population level (*23*). The discovery that plasma of COVID-convalescent individuals contains polyclonal Ig with CoV2-neutralizing activity (*24*) paved the way for multiple clinical trials testing the efficacy of convalescent plasma therapy against COVID (*25*). Whether convalescent plasma therapy was efficacious remains debated (*26*). What is not debated, however, is the efficacy of vaccines which were designed to elicit Ig against CoV2. In the US, the first COVID vaccines available comprised either a two-dose encapsulated mRNA formulation (BNT162b2 or mRNA-1273) or a single-dose adenovirus vector formulation (Ad26.COV2.S). The US Food & Drug Administration (FDA) granted emergency use authorizations (EUA) for BNT162b2 and mRNA-1273 on Dec 11 2020 and Dec 18 2020, respectively (*27, 28*); the FDA EUA for Ad26.COV2.S was granted on Feb 27 2021 (*29*). The advent of these and other COVID vaccines led to dramatic declines in COVID morbidity and mortality (*30*), and—relative to vaccinated individuals—unvaccinated individuals are more likely to need hospitalization or die following CoV2 infection (*31*).

Since interrupting the Spike:ACE2 interaction was the goal of now-approved vaccines (*32, 33*), and remains a goal of potential COVID therapies (*34, 35*), the continual emergence of new CoV2 lineages with numerous and diverse Spike mutations threatens our ability to prevent and treat future CoV2 infections. It is therefore important to understand the relationships between CoV2 lineage emergence, CoV2-specific Ig levels—as elicited by either natural infection or vaccination—and their neutralization capacity. This is especially true of asymptomatic individuals who are PCR positive (PCR^POS^), as they are estimated to account for 50-65% of all transmission (*36, 37*). Here, we describe the relationships between COVID incidence, CoV2 lineage, viral load, CoV2-specific Ig responses (IgM, IgA & IgG) and inhibitory capacity in the saliva of asymptomatic PCR^POS^ individuals, as the oral cavity and saliva—in addition to being readily accessible—are important sites of CoV2 infection and transmission (*38*) (especially newer Omicron variants (*39-43*)). CoV2-specific Ig responses were similarly assessed in PCR^NEG^ individuals with a history of CoV2 infection and/or COVID vaccination with pre-Omicron vaccines. These data were generated as part of a large university COVID monitoring program which occurred between Aug 2020 → Jun 2022.

## METHODS

### Institutional approval statement

This work was reviewed and approved by The Ohio State University Biomedical Sciences Institutional Review Board (ID #2021H0080). This work was also reviewed and approved by the Ohio State Institutional Biosafety Committee (IBC) (ID #2020R00000046).

### Saliva specimen collection and handling

The Ohio State COVID monitoring program was active from Aug 2020 through June 2022. As part of this program, saliva specimens were collected on a weekly basis from students, staff and faculty who self-reported as being asymptomatic at the time of specimen collection. On and prior to the day of saliva collection at one of several mass testing sites (**FIG 1A**), individuals were instructed to define themselves *symptomatic* if they had at least one or more of the following : fever, chills, shortness of breath, difficulty breathing, fatigue, muscle aches, body aches, headache, new loss of taste, new loss of smell, sore throat, congestion, runny nose, nausea, vomiting, or diarrhea. To prevent contagion, symptomatic individuals were instructed not to come to the mass testing site and were instead referred to a healthcare provider for follow-up (e.g. the campus student health clinic). Individuals were defined as *asymptomatic* if they had none of the symptomatic conditions listed above. On the day of testing, individuals were instructed to refrain from food or drink for 30 minutes prior to collection, and to gently eject saliva into the collection tube, swallowing first and keeping saliva free from mucus, until the 1 mL mark on a sterile conical was reached (i.e. passive drool method). Specimens from asymptomatic individuals were collected at each of the six Ohio State campuses in Franklin county (OSU-Columbus), Licking county (OSU-Newark), Richland county (OSU-Mansfield), Allen county (OSU-Lima), Marion county (OSU-Marion) and Wayne county (OSU-Wooster). Specimens were then couriered to the CLIA-approved Applied Microbiology Services Lab (AMSL) of the Ohio State Infectious Disease Institute (IDI) and analyzed in accordance with the SalivaDirect assay, a clinical diagnostic test that is Emergency Use Authorization (EUA) approved by the US Food & Drug Administration (FDA) for SARS-COV-2 detection (*44*). While performing the SalivaDirect real time polymerase chain reaction (PCR), saliva samples were stored in a 4°C cold room until they were deemed either PCR negative (PCR^NEG^) or PCR positive (PCR^POS^) for CoV2. Per the SalivaDirect method (*45*), any sample with a C_T_ value ≤ 40 was considered PCR^POS^ for CoV2. The positive or negative status of the sample was reported to the individual and regional public health authorities (Columbus Public Health, Ohio Department of Health, ODH,) per state and federal policies at the time. PCR^POS^ saliva samples and select PCR^NEG^ saliva samples were then removed from the 4°C cold room, aliquoted into microcentrifuge tubes, frozen (−20°C) and analyzed for viral genome sequencing and lineage identification, as well as host antibody response characterization.

**FIGURE 1.**
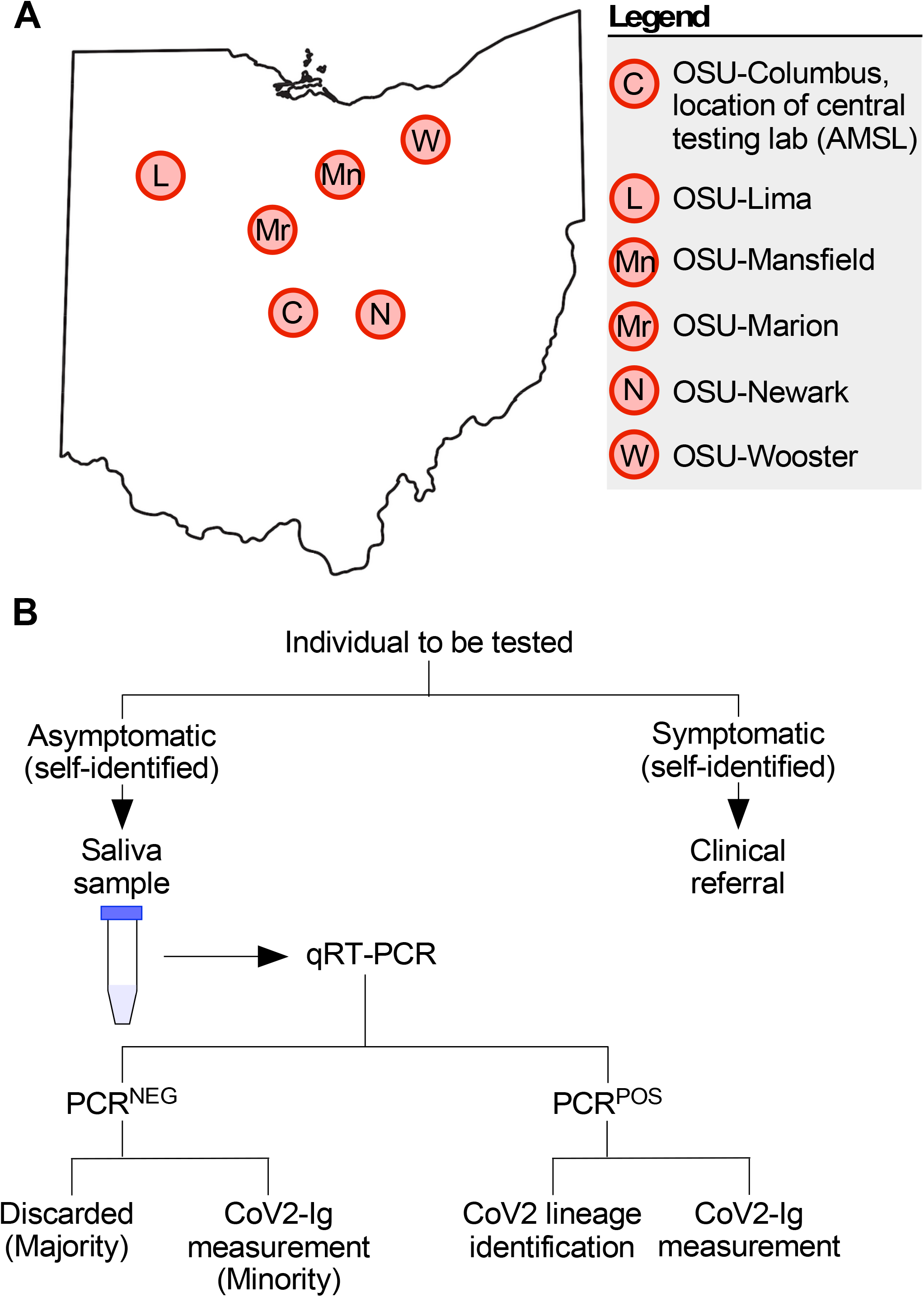
Overview of our university COVID monitoring program and workflow. (**A**) Map of Ohio with locations of the six university campuses which participated in the COVID monitoring program. (**B**) On and prior to the day of testing, each individual assessed themselves for one or more COVID symptoms (see *Methods* for complete list). If symptomatic, the individual was given a clinical referral and instructed to not go to their on-campus testing facility, to prevent contagion. If asymptomatic, the individual provided a saliva sample which was tested (typically within 24 hours of sample provision) via qRT-PCR for the presence of the CoV2 N gene. Individuals were notified as soon as possible as to whether their sample was negative (PCR^NEG^) or positive (PCR^POS^) for the virus, a positive result being a C_T_ ≤ 40. PCR^POS^ samples were subsequently aliquoted and used for both CoV2 lineage identification and measuring the concentrations of immunoglobulin against specific CoV2 antigens (CoV2-Ig). The vast majority of PCR^NEG^ samples were discarded; however, a minority were retained and used for CoV2-Ig measurements. PCR^POS^ and PCR^NEG^ samples were otherwise treated identically.

### Sequencing and Lineage Identification

PCR^POS^ saliva samples with a C_T_ ≤ 33 had their whole CoV2 viral genome sequenced and lineage assigned per the methods described in our previous work (*46*) (samples with a C_T_ > 33 had insufficient viral RNA for sequencing). CoV2 genome copy numbers were calculated via linear regression analysis, by comparison to the C_T_ values of SalivaDirect reference standards. CoV2 genome sequences were submitted to the Global Initiative on Sharing Avian Influenza Data (GISAID) database in a manner consistent with ODH expectations and policies at that time, in as close to real time as possible. The abbreviations we use for each lineage in this study and associated figures are as follows: CoV2^Anc^, the ancestral lineage of CoV2 which emerged from Wuhan, China; CoV2^US^, the B.1.2 lineage which was among the first detected in our region of the United States (*46-48*); CoV2^Alpha^, the B.1.1.7 lineage or Alpha variant of concern (VOC) which was first reported by the UK in Dec 2020 (*3*); CoV2^Beta^, the B.1.351 lineage or Beta VOC which was first reported in South Africa in Dec 2020 (*4*); CoV2^Gamma^, the P.1 lineage or Gamma VOC which was first reported in Brazil in Jan 2021 (*9*); CoV2^Delta^, the B.1.617.2 lineage or Delta VOC which was first reported in India in Dec 2020 (*10*); CoV2^Omicron^, the B.1.1.529 lineage or Omicron VOC which as first reported in South Africa in Nov 2021 (*11*); CoV2^O-BA.1^, the BA.1 variant of CoV2^Omicron^; CoV2^O-BA.2^, the BA.2 variant of CoV2^Omicron^; CoV2^O-BA.4^, the BA.4 variant of CoV2^Omicron^; CoV2^O-BA.5^, the BA.2 variant of CoV2^Omicron^. The nonsynonymous Spike mutations which distinguish these lineages are depicted in supplemental **FIG S1**. Any lineage which was not a VOC or otherwise not mentioned above (e.g. Epsilon) is labeled “Non-VOC.”

### COVID wave designations and comparisons

We defined a COVID wave within our university community as when new PCR^POS^ case counts rose above the overall period median for ≥ 3 weeks in a row (the overall period being Jan 2021 through Jun 2022). For comparisons to COVID incidence in surrounding counties, we accessed publicly available ODH data via their public-facing dashboard (accessed Nov 14 2022).

### Measuring Binding Antibody Levels in Saliva

After PCR results were reported (typically within 24 hours of specimen collection), PCR^POS^ and select PCR^NEG^ specimens were removed from the 4°C cold room, aliquoted into microcentrifuge tubes containing Triton X-100 to inactivate CoV2 (final concentration: 1% Triton X-100) (*49*). PCR^NEG^ samples were selected based on the donors’ having had either a prior CoV2 infection (allowing us to measure durability of the antibody response following natural infection) or their having been vaccinated against COVID (allowing us to compare the antibody responses of uninfected vaccinated individuals to those of infected vaccinated individuals, a.k.a. breakthrough infections). All samples were treated identically regardless of whether they were PCR^POS^ or PCR^NEG^. Following the addition of Triton X-100, samples were vortexed and allowed to incubate for 1 hour at room temperature (*49*). Samples were subsequently stored at -80°C until the antibody levels in all samples could be measured at the same time, thus eliminating batch effects. The Meso Scale Diagnostics (MSD) V-Plex platform was used to measure the concentration of CoV2 antigen specific immunoglobulin (IgM, IgA and/or IgG) in PCR^POS^ and PCR^NEG^ samples. Briefly, the MSD V-Plex assay comprises a 96-well plate which, within each well, contains multiple spots that were coated with defined antigens. For our study, these antigens included recombinant forms of three CoV2^Anc^ lineage proteins (Nucleocapsid [N], Spike, and the Spike Receptor Binding Domain [RBD]), as well as CoV2^Alpha^ Spike, CoV2^Beta^ Spike, CoV2^Gamma^ Spike, and CoV2^Delta^ Spike (**FIG S1**). The Spike antigens consisted of the trimerized form of the ectodomain; the N antigen consisted of the full-length protein. Antibodies in the sample bind to the antigens, and reporter-conjugated secondary antibodies were used for detection. Saliva samples were thawed on ice and diluted by a factor of 10 in the diluent provided in the V-Plex assay kit for each assay. The V-Plex assays were performed according to manufacturer instructions, and plates were read on an MSD instrument which measures light emitted from reporter-conjugated secondary antibodies. Using MSD’s analysis software, the light signal measured by the MSD instrument was converted into arbitrary units (AU) representing amount of antibody present relative to the standard curve of the assay. The AU values for IgM, IgA and IgG binding to CoV2^Anc^ N, Spike, and Spike RBD were transformed to WHO binding antibody units (BAU) via validated WHO standards and conversion factors provided by MSD. The AU values for IgM, IgA and IgG binding to other forms of N or Spike (i.e., those of VOC) cannot be converted to WHO BAU, as there are no WHO standards for these recombinant proteins. For this reason, the levels of each Ig isotype which bind to CoV2^Alpha^, CoV2^Beta^, CoV2^Gamma^, and CoV2^Delta^ forms of Spike are expressed as AU.

### Spike inhibition assay

The capacity of saliva specimens to inhibit Spike activity was quantified using a commercially available ACE2 displacement assay (MSD COVID-19 ACE2 Neutralization Kit method). Plate-bound Spike was incubated with diluted saliva (the same specimens used for Ig measurements) per manufacturer protocols, followed by washing and addition of a luminescent probe-conjugated, recombinant form of human ACE2. The extent to which luminescence declined relative to non-saliva (i.e., diluent only) treated wells was used to derive a percent inhibition value for each individual sample, using the following formula: % inhibition = 1 – (saliva sample luminescence value / diluent only luminescence value) × 100.

### Graphing and statistics

Graphs were generated in RStudio or GraphPad. All statistical tests were performed in RStudio. Data was tested for normality using the Kolmogorov-Smirnov test and for equal variance using the Bartlett test of homogeneity of variances. For data that did not have normal distribution, the Kruskal-Wallis rank sum test was used to determine if there were significant differences between groups in unpaired datasets, and the Friedman rank sum test was used in paired datasets. Within those datasets, the significant differences between groups were identified via an unpaired or paired Wilcoxon rank sum test as appropriate with Benjamini-Hochberg p value adjustment method. For the neutralization data which contained several zero values, the Shapiro-Wilk normality test was used, followed by the Bartlett test of homogeneity of variances. The Kruskal-Wallis rank sum test was used to determine if significant differences were present, followed by Dunn’s test with Benjamini-Hochberg p value adjustment method to identify which groups were significantly different. Differences between groups were considered significant if *P* < 0.05 and are graphically indicated by 1 or more asterisks (\**P* < 0.05; \*\**P* < 0.005; \*\*\**P* < 0.0005).

## Data Availability

All data produced in the present work are contained in the manuscript

## Abbreviations

The following abbreviations are used throughout our manuscript:

PCR^POS^: an individual or saliva specimen that was PCR positive for CoV2 (C_T_ value ≤ 40) ;
PCR^NEG^: an individual or saliva specimen that was PCR negative for CoV2;
Spike and N: unless otherwise stated the Spike and N proteins of CoV2 (not any other coronavirus);
CoV2-Ig: immunoglobulin of any isotype that recognizes any CoV2 antigen;
IgM^Spike^: IgM that recognizes Spike;
IgA^Spike^: IgA that recognizes Spike;
IgG^Spike^: IgG that recognizes Spike;
IgG^RBD^: IgG that recognizes the Spike Receptor Binding Domain;
IgG^N^: IgG that recognizes the N protein;
Vax^POS^: an individual who was fully vaccinated against COVID (but not boosted) prior to saliva specimen collection;
Vax^NEG^: an individual who was not fully vaccinated against COVID prior to saliva specimen collection;
New^POS^: an individual who at the time of saliva collection was PCR^POS^ for the first time;
Prior^POS^: an individual who at the time of saliva collection was PCR^NEG^ but who had a prior CoV2 infection (i.e. the individual had been PCR^POS^ 3-9 months prior).

## RESULTS

### I. Study overview

The first confirmed cases of COVID in the state of Ohio were reported on Mar 9 2020 (*50*). The Ohio State University suspended on campus activities the same day (*51*) and subsequently developed a campus wide plan to monitor the incidence of CoV2 infection among its students, staff and faculty (*52*). Individuals participating in this monitoring program, which formally began in Aug 2020, provided saliva on a weekly basis for COVID testing. Prior to testing, individuals who self-reported as being symptomatic were not tested and were instead given a clinical referral (see *Methods* for additional details). Individuals who self-reported as being asymptomatic provided a saliva specimen via a passive drool method at each of our six university campuses (**FIG 1A**). Specimens were assessed by our CLIA-certified lab for the presence of CoV2 using real-time quantitative reverse transcription PCR (qRT-PCR). Specimens were not pooled prior to testing. qRT-PCR results were reported to the individual and the regional public health authority per state and federal policies at the time. If a specimen had a C_T_ value ≤ 40 it was considered positive for CoV2 virus (PCR^POS^). Per our Institutional Review Board (IRB) approved protocols and workflows (**FIG 1B**), PCR^POS^ saliva samples were subsequently used for CoV2 lineage identification and CoV2-specific immunoglobulin (CoV2-Ig) measurements. In some instances, select saliva samples that were negative for CoV2 virus (PCR^NEG^) were also collected, the reasons for which will be made clear in sections below. The relationships between these molecular and immunological readouts to one another, as well as to coded data concerning the prior infection status and vaccination status of the saliva donor, are described below for the period spanning Jan 2021 (before COVID vaccines were widely available to students in our university community) to June 2022, when the monitoring program ended. See Methods for details regarding saliva collection, symptomatic versus asymptomatic designation, qRT-PCR, CoV2 lineage identification, CoV2-Ig measurements, and statistical analyses. In total, >850,000 diagnostic PCR tests were performed by our lab during this monitoring program.

### II. The incidence of CoV2 positivity in our university community occurred in waves which reflected those occurring in surrounding regions

The incidence of new PCR^POS^ cases among asymptomatic individuals in our university community, for the period spanning Jan 2021→June 2022, is shown in **FIG 2**. COVID monitoring occurred before Jan 2021; however, because the bulk of PCR testing at that time was contracted to a commercial entity, our access to the raw PCR data before Jan 2021 is limited. Above these data are two timelines relevant to data interpretation, indicating when Ohio COVID vaccination policies shifted from prioritizing at risk populations (e.g. elderly) to anyone ≥16 years of ages well as the deadlines for all our community members (i.e. university students, faculty and staff) to have received their first and second COVID doses (Oct 15 2021 and Nov 15 2021, respectively) (*53*). Indicated below the data are corresponding intervals in the academic calendar, which will be referred to in subsequent sections. We identified 11,989 PCR^POS^ individuals between Jan 2021→June 2022; the median, mean and maximum new PCR^POS^ cases per test day were 15, 34 and 523, respectively. There were, however, six time periods when the new case counts rose above the overall period median for ≥ 3 weeks in a row. These six time periods are hereafter referred to as Waves 1–6 and spanned the following dates: *Wave 1*, Jan 11 2021→ Jan 29 2021; *Wave 2*, Feb 22 2021→ Mar 12 2021; *Wave 3*, Mar 22 2021→ Apr 16 2021; *Wave 4*, Aug 16 2021→ Sep 24 2021; *Wave 5*, Nov 15 2021→ Feb 18 2022; *Wave 6*, Apr 18 2022→ May 6 2022. The waves of COVID incidence amongst asymptomatic individuals in our university community mirrored (rather than preceded) the waves of COVID incidence in the counties surrounding each university campus (*54*) (supplemental **FIG S2**).

**FIGURE 2.**
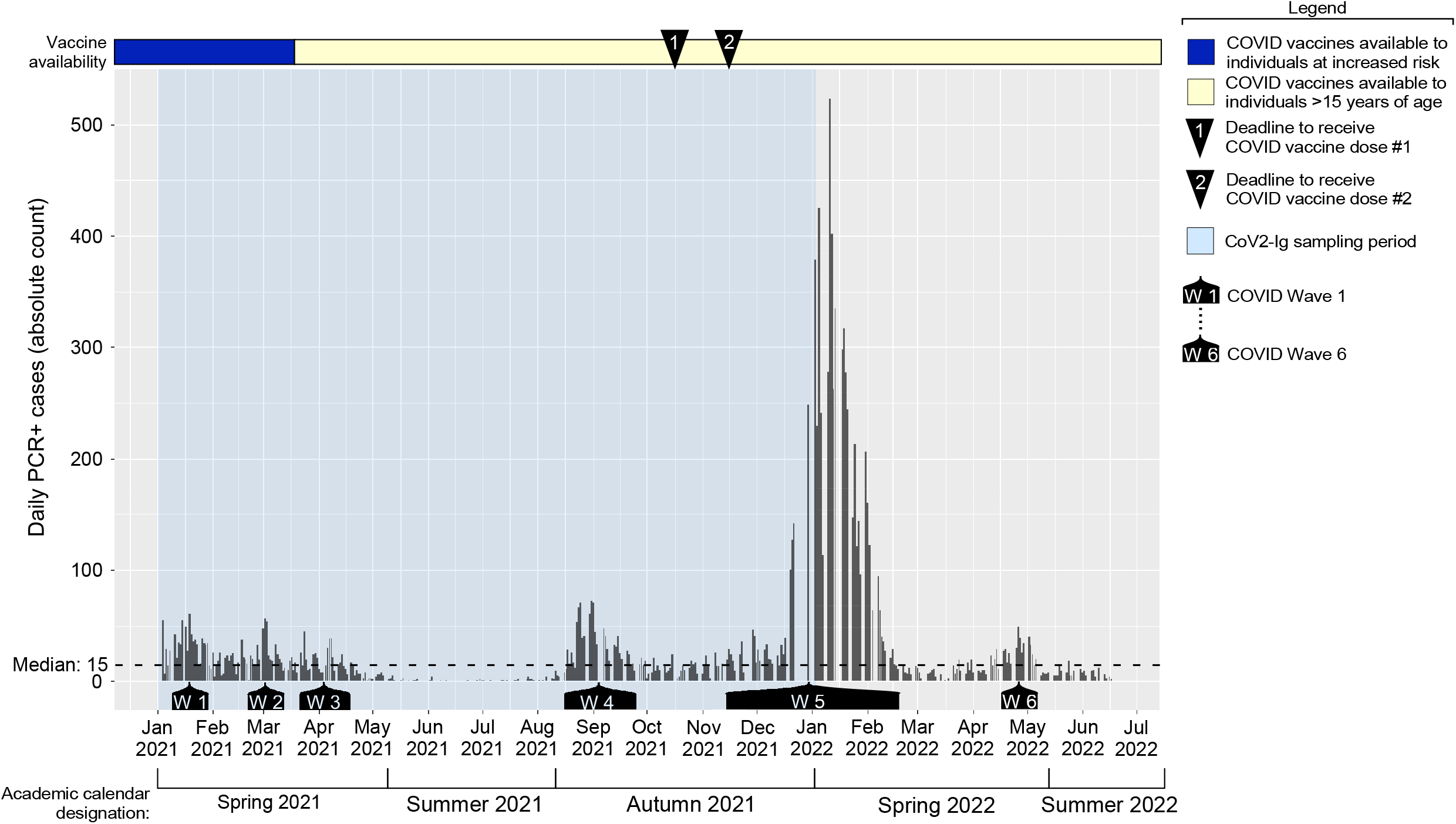
The incidence of PCR positivity among asymptomatic members of our university community. Saliva samples from asymptomatic individuals were collected on a daily basis and tested by qRT-PCR for the presence of the CoV2 N gene. Shown are the number of PCR^POS^ saliva samples identified each day during the period spanning Jan 2021 → May 2022, with each bar representing a single day. Above the graph is a timeline depicting when COVID vaccine availability shifted in Ohio (i.e. when the national vaccination priority expanded from vulnerable populations to encompass anyone >15 years of age), as well as indications of the deadlines by which all university community members were required to have received their first and second vaccine dose of either the BNT162b2 or mRNA-1273 vaccines. Below the graph are indications of the periods we refer to as Waves 1-6, a wave being defined as when the daily PCR^POS^ case count exceeded the period median (15) for ≥3 weeks, as well Blue shading indicates when the samples we used for CoV2 Ig measurements were collected.

### III. Prior to community vaccine requirements being established, CoV2 was becoming progressively more concentrated in the saliva of asymptomatic individuals

The emergence of CoV2 lineage variants in multiple Ohio communities (*55-61*) with potential for greater infectivity and/or transmissibility led us to assess the relationship between CoV2 abundance in saliva and variant identity. We used the qRT-PCR cycle threshold (C_T_) value as a readout of CoV2 abundance, as the SalivaDirect C_T_ value is inversely proportional to CoV2 viral load (i.e. a lower C_T_ value corresponds to higher CoV2 RNA levels in the tested sample) (*45*), and a commonly used as a proxy for probability of transmission (i.e. a lower C_T_ value correspond to higher transmission probability) (*62-66*). Variant identity was determined by next generation sequencing of the entire CoV2 genome and subsequent alignment with Global Initiative on Sharing Avian Influenza Data (GISAID) reference sequences. During the entire monitoring period, CoV2 genome sequences were submitted to the GISAID database in a manner consistent with ODH expectations and policies at that time, in as close to real time as possible.

The weekly composite and daily individual C_T_ values of each PCR^POS^ individual over this period are shown in **FIG 3A** and **FIG 3B**, respectively, with color annotations in **FIG 3B** indicating the lineage identity. The same data are presented in two ways (weekly composite versus daily individual) in order to best illustrate the following trends: During Wave 1 (Week 2 of Jan 2021 → Week 4 of Jan 2021), the median weekly C_T_ values of all PCR^POS^ saliva samples ranged between 29 - 30.5 (**FIG 3A**). During Wave 2 (Week 4 of Feb 2021 → Week 2 of Mar 2021) the median weekly C_T_ values ranged between 28 - 30.5 (**FIG 3A**). The median C_T_ range lowered and tightened during Wave 3 (Week 4 of Mar 2021 → Week 2 of Apr 2021) median weekly C_T_ values ranged from 28.5 - 29.5 (**FIG 3A**). The number of tests performed fell precipitously during Jun 2021 and Jul 2021, as the campus population is minimal during the summer months; therefore, we are reluctant to draw conclusions from or otherwise compare Summer 2021 C_T_ value data to the prior semester, when testing volume was higher. Upon resumption of high-volume testing during the later weeks of August 2021, which marked the beginning of the Autumn 2021 semester and Wave 4 (Week 3 of Aug 2021 → Week 3 of Sept 2021), we noted the lowest median C_T_ range of all waves (27.0 - 27.5) (**FIG 3A**). The cumulative C_T_ data during each wave (**FIG 3C**) and extrapolated CoV2 genome copy concentrations (**FIG 3D**) are consistent with Wave 4 saliva samples having the highest virus concentrations of all waves. Wave 5 was the longest wave (Week 3 of Nov 2021 → Week 2 of Feb 2022) with daily PCR^POS^ cases reaching a maximum of 523 on Jan 11, 2022. The C_T_ value range during Wave 5, which followed our community deadline for vaccine requirements, was significantly higher than that of Wave 4 (**FIG 3B**). The last wave before the COVID monitoring program ended, Wave 6 (Week 2 of Apr 2022 → Week 1 of May 2022), had lower C_T_ values than Wave 5 (**FIG 3B**). The lowest C_T_ value we ever observed was on Feb 18 2021 (C_T_=14.2).

**FIGURE 3.**
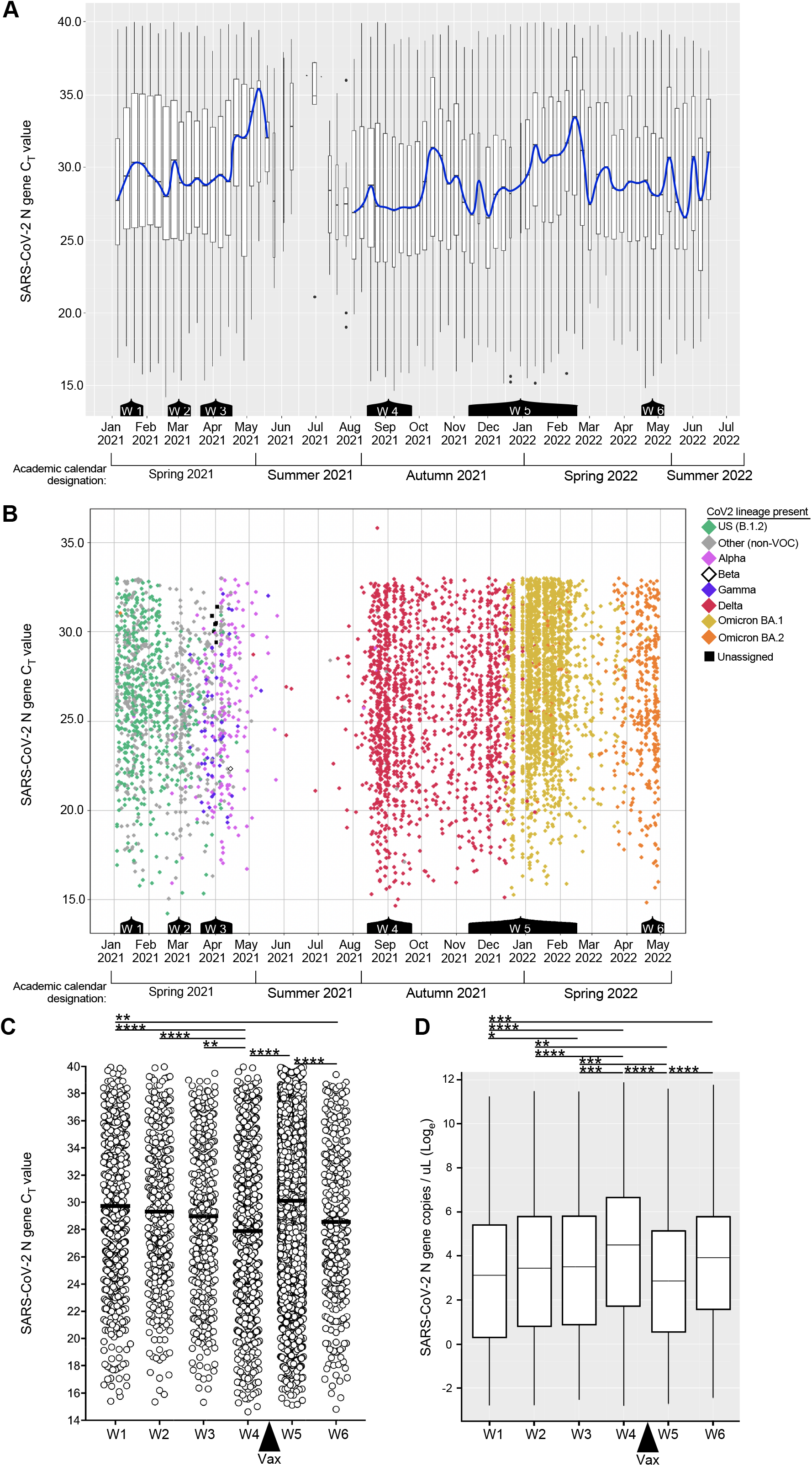
Saliva CoV2 viral loads among asymptomatic members of our university community. (**A**) Box plot representation of all the C_T_ values of all the PCR^POS^ saliva samples during each week of the period spanning Jan 2021 → Jun 2022. The blue line passes through the median C_T_ value of each week. Below the graph are indications of the periods corresponding to Waves 1-6 of the prior figure. (**B**) Scatter plot representation of the same C_T_ value data as in (**A**) above, the exceptions being daily data are shown (as opposed to weekly composites) and samples with a C_T_ >33 are omitted (these could not be sequenced due to insufficient amounts of genetic material). Each dot represents an individual sample; the color of each diamond indicates the CoV2 lineage present (Green, CoV2^US^; Pink, CoV2^Alpha^; White, CoV2^Beta^ ; Blue, CoV2^Gamma^; Red, CoV2^Delta^; Gold, CoV2^O-BA.1^; Orange, CoV2^O-BA.2^). Gray diamonds indicate samples whose lineage was not a VOC. Black squares indicate a sequence that did not align to known lineages and thus could not be assigned. Note that CoV2^Beta^ only appeared once in our university community, on Apr 15 2021. (**C**) The C_T_ value and (**D**) calculated CoV2 genome copy concentration in of each positive sample during Wave 1, Wave 2, Wave 3, Wave 4, Wave 5 and Wave 6. The “Vax” arrow indicates when community vaccine requirements went into effect (after Wave 4, before Wave 5). Asterisks indicate those inter-wave differences that were statistically significant, as determined by one way ANOVA (* p≤ 0.05, ** p≤ 0.005, *** p≤ 0.0005, *** p≤ 0.00005).

### IV. Each wave of CoV2 positivity corresponded to the emergence of a new CoV2 lineage within our community

The CoV2 lineages present in each individual PCR^POS^ sample during the same time periods as above are shown in **FIG 3B**, exceptions being samples with a C_T_ value of >33 as these could not be sequenced due to the viral RNA levels being too low. Among sequenced samples, the median and mean ages of infected individuals were 21 and 23, respectively, and varied minimally during the monitoring period (supplemental **FIG S3**). Males were more likely to meet sequencing criteria (i.e. a C_T_ ≤ 33) than females during Waves 1-3 (supplemental **FIG S4**); this was not true of later Waves, however, and female representation was higher during the entire monitoring period overall (supplemental **FIG S4**). During the period spanning Jan 2021 to mid-Feb 2021, the predominant lineage was B.1.2, which we hereafter refer to as CoV2^US^ since it was among the first detected in our region of the United States (*46-48*). The period of CoV2^US^ lineage predominance corresponds to Wave 1 in our community (**FIG 2, FIG 3B**). Beginning mid-Feb 2021 and extending to mid-Mar 2021 was a period of time when an array of lineages which we collectively refer to as “non-VOC” were predominant, as they were more diverse compared to earlier and later testing periods and were never considered to be Variants of Concern (VOC). Although the CoV2^US^ lineage was still being detected, Wave 2 primarily comprised of non-VOCs (**FIG 3B**). As the Ides of March approached in 2021, so too did two VOCs begin appearing with increasing frequency: the Alpha VOC (CoV2^Alpha^) and Gamma VOCs (CoV2^Gamma^). CoV2^Alpha^ and CoV2^Gamma^ were widely considered at that time to be more transmissible than previous lineages (*9, 67*). CoV2^Alpha^ and CoV2^Gamma^ were the primary lineages detected during Wave 3 (**FIG 3B**), and continued to predominate among the few positive samples collected during May 2021. The Beta VOC (CoV2^Beta^) only appeared once in our university community (Apr 15, 2021). Beginning Jun 2021 and continuing through Dec 2021 the new Delta VOC (CoV2^Delta^) made up the vast majority of PCR^POS^ saliva samples (**FIG 3B**). CoV2^Delta^ is more transmissible than CoV2^Alpha^ and CoV2^Gamma^ (*68*), and the period in which CoV2^Delta^ predominated coincided with COVID Wave 4 in our community (**FIG 2**). Wave 5, the penultimate and largest COVID wave, coincided with the emergence and dominance of the Omicron VOC (CoV2^Omicron^) subvariant, BA.1 (CoV2^O-BA.1^). Wave 6, final wave before our COVID monitoring program ended, was dominated by the CoV2^Omicron^ subvariant BA.2 (CoV2^O-BA.2^) (**FIG 3B**). When considered alongside the C_T_ values and CoV2 genome copy numbers that characterized each wave (**FIG 3C-D**), the above data demonstrate that the shift from CoV2^US^ → CoV2^Alpha^/CoV2^Gamma^ → CoV2^Delta^ coincided with the virus becoming progressively more concentrated in the saliva of asymptomatic individuals, this trend ending after community vaccine requirements were established.

### V. Among pre-Omicron lineages, CoV2^Delta^ elicited the highest levels of Spike-specific IgA and IgG in unvaccinated, asymptomatic individuals

In symptomatic CoV2 PCR^POS^ individuals, CoV2-specific Ig levels in the circulation and airways increase at variable rates depending on the isotype (*22*). To assess whether CoV2-specific Ig was detectable in the saliva of asymptomatic CoV2 PCR^POS^ individuals, as well as whether levels of the same Ig varied depending on the CoV2 lineage present, we used the same samples described above (i.e. those used for lineage identification) to measure saliva levels of CoV2 Spike-specific IgM (IgM^Spike^), CoV2 Spike-specific IgA (IgA^Spike^) and CoV2 Spike-specific IgG (IgG^Spike^) (**FIG 4**). Individuals vaccinated against COVID were excluded from this analysis (the vaccination record of each person in our university community was closely monitored during this time period), and saliva samples from individuals infected with CoV2^Anc^, CoV2^Alpha^ and CoV2^Gamma^ were collected prior to COVID vaccines being widely available in our community; therefore, no vaccine-elicited antibody responses would be expected in these samples. Among individuals infected with CoV2^Delta^, only unvaccinated individuals were included in the **FIG 4** analysis. To eliminate viral load as a confounding variable, only PCR^POS^ saliva samples with similar C_T_ range were used for Ig comparisons (C_T_ range = 22-26). PCR^NEG^ saliva collected in early 2020 from healthy individuals living in the US and no COVID history were used to estimate “pre-pandemic” levels of IgM^Spike^, IgA^Spike^ of IgG^Spike^ binding.

**FIGURE 4.**
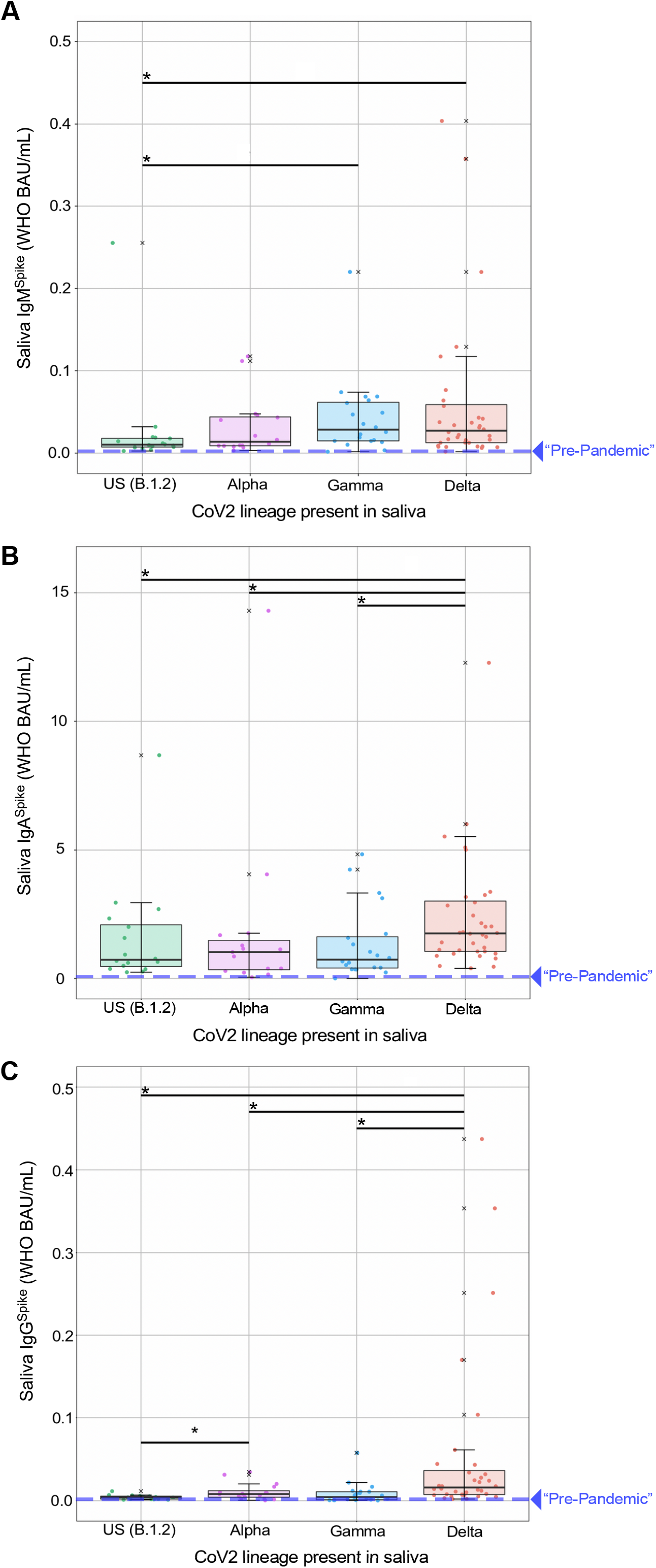
Spike-specific Ig levels in the saliva of newly positive, asymptomatic individuals at the time of PCR testing. Saliva samples from individuals who were newly positive (New^POS^, PCR positive for the first time ever) for either the CoV2^US^, CoV2^Alpha^, CoV2^Gamma^ or CoV2^Delta^ lineage were used to measure the concentrations of (**A**) IgM^Spike^, (**B**) IgA^Spike^ and (**C**) IgG^Spike^. The CoV2^Anc^ Spike was used as the capture antigen in each case, and concentrations are expressed as World Health Organization (WHO) binding antibody units (BAU) per mL. PCR^NEG^ saliva collected in early 2020, from healthy individuals living in the US with no COVID history, was tested in the same manner used to estimate “pre-pandemic” levels of IgM^Spike^, IgA^Spike^ and IgG^Spike^ binding, which are represented by the dashed lines on each graph. ^X^, values that were considered outliers but are nevertheless shown for completeness and are included in all statistical group comparisons. * p≤ 0.05, as determined by unpaired Wilcoxon tests with Benjamini-Hochberg adjustment.

Saliva IgM^Spike^ (**FIG 4A**), IgA^Spike^ (**FIG 4B**) and IgG^Spike^ (**FIG 4C**) data are shown relative to which CoV2 lineage was detected in the same saliva donor (CoV2^US^, CoV2^Alpha^, CoV2^Gamma^ or CoV2^Delta^) and are expressed as WHO binding antibody units, or BAUs. As shown in **FIG 4A-C**, respectively, nearly all PCR^POS^ individuals had saliva IgM^Spike^, IgA^Spike^ and IgG^Spike^ levels that were above “pre-pandemic” levels, regardless of whether they were infected with CoV2^US^, CoV2^Alpha^, CoV2^Gamma^ or CoV2^Delta^. There were, however, three noteworthy differences between PCR^POS^ individuals depending on the lineage present. First, whereas individuals infected with CoV2^US^ and CoV2^Alpha^ had similar IgM^Spike^ levels, those infected with CoV2^Gamma^ and CoV2^Delta^ had higher IgM^Spike^ levels relative to those infected with CoV2^US^ (**FIG 4A**). Second, saliva IgA^Spike^ levels were similar between individuals infected with CoV2^US^, CoV2^Alpha^ and CoV2^Gamma^; CoV2^Delta^ infected individuals, on the other hand, had significantly higher IgA^Spike^ levels compared to those infected with CoV2^US^, CoV2^Alpha^ or CoV2^Gamma^ (**FIG 4B**). Third, saliva IgG^Spike^ levels were elevated in CoV2^Alpha^-infected individuals relative to CoV2^US^-infected individuals (**FIG 4C**); however and analogous to IgA^Spike^ differences (**FIG 4B**), CoV2^Delta^-infected individuals had significantly higher IgG^Spike^ levels compared to those infected with CoV2^US^, CoV2^Alpha^ or CoV2^Gamma^ (**FIG 4C)**. For IgM^Spike^, IgA^Spike^ and IgG^Spike^ measurements, the recombinant Spike antigen used for Ig detection was identical to that of CoV2^Anc^, as this enabled data transformation to WHO BAU (see *Methods*); the same patterns were observed, however, when the same saliva samples were tested against recombinant CoV2^Alpha^, CoV2^Beta^, and CoV2^Gamma^ Spike antigens (**Supplemental FIG S5)**.

### VI. Following infection of unvaccinated individuals, IgG^Spike^ and IgG^RBD^ persisted at higher levels in saliva than IgG^N^

To determine the extent to which CoV2-specific IgG in saliva was sustained over time, we performed the analysis shown in **FIG 5** wherein saliva IgG^Spike^ levels, as well as Nucleocapsid (N)-specific IgG (IgG^N^) levels, were compared across two groups of individuals: “New^POS^” individuals who, at the time of saliva collection, were positive for either CoV2^US^ or CoV2^Alpha^; “Prior^POS^” individuals who were uninfected at the time of saliva collection, but had been PCR^POS^ 3-9 months earlier. In this instance, saliva samples from Prior^POS^ individuals were collected in May 2021. Most individuals in our Prior^POS^ cohort were infected during the Autumn 2020 semester, before COVID vaccines were available; therefore, any saliva IgG^Spike^ or IgG^N^ present would have formed after natural infection (not vaccination). As in the previous figure, individuals with a record of COVID vaccination were excluded from this analysis.

**FIGURE 5.**
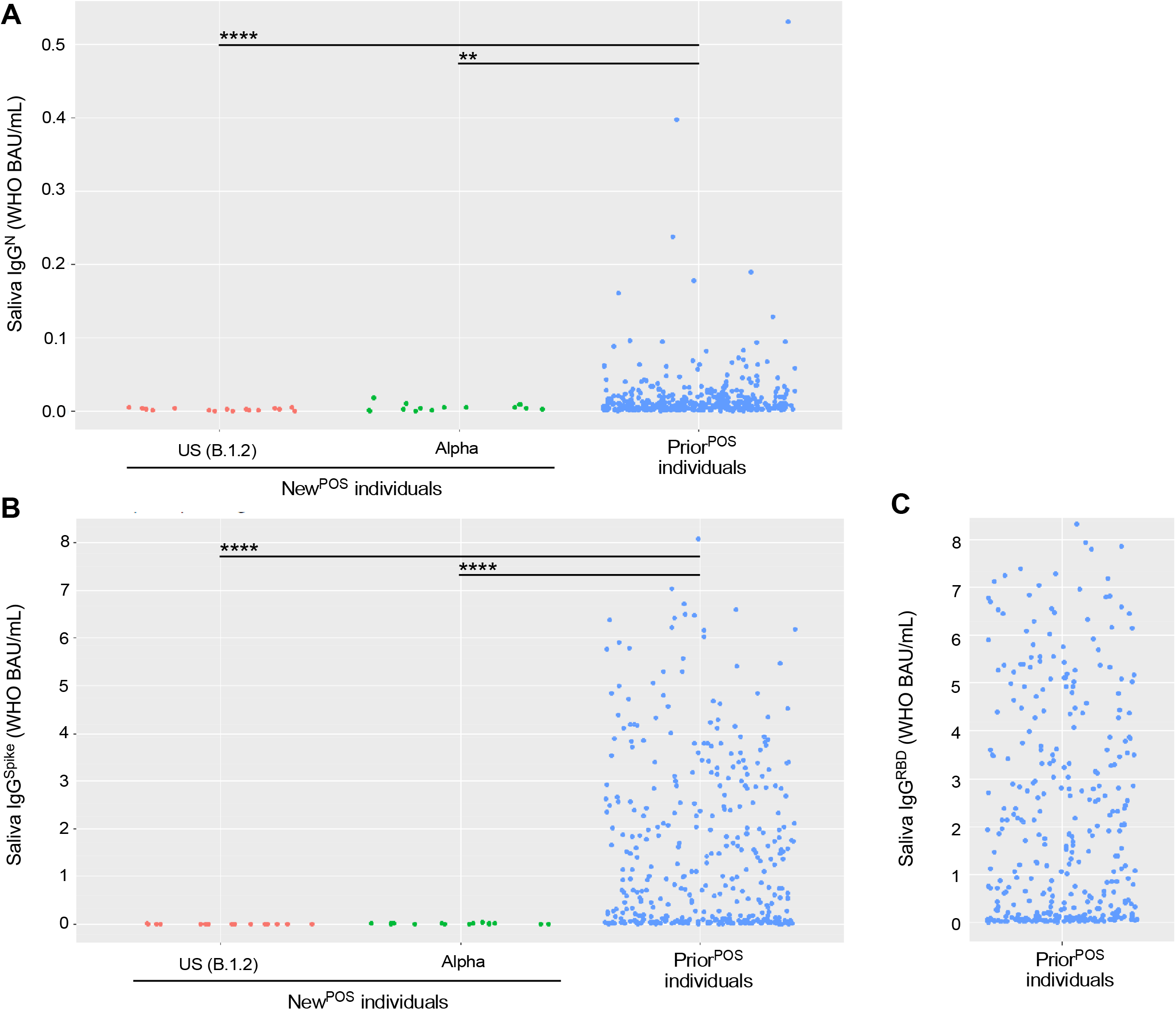
Nucleocapsid- and Spike-specific IgG levels in saliva of newly positive, asymptomatic individuals versus prior positive, asymptomatic individuals. Saliva from New^POS^ individuals infected with either CoV2^US^ or CoV2^Alpha^, as well as PCR^NEG^ saliva from individuals who had been infected 3-9 months prior (Prior^POS^) with either CoV2^US^, CoV2^Alpha^ or a non-VOC, were used to measure the concentrations of (**A**) Nucleocapsid-specific IgG, (**B**) Spike-specific IgG, and (**C**) Spike RBD-specific IgG. * p≤ 0.05, as determined by unpaired Wilcoxon test with Benjamini-Hochberg adjustment.

Among New^POS^ individuals, saliva IgG^N^ levels were similar regardless of whether they were infected with CoV2^US^ or CoV2^Alpha^ (**FIG 5A**), as were saliva IgG^Spike^ levels (**FIG 5B;** note the data points in the first two columns of **FIG 5B** are the same as those in the first two columns of **FIG 4C**). Relative to New^POS^ individuals, saliva IgG^N^ levels in Prior^POS^ individuals were higher (**FIG 5A**); however, the difference in saliva IgG^Spike^ levels between New^POS^ versus Prior^POS^ individuals was more pronounced (**FIG 5B**). Saliva IgG^Spike^ levels were highest in Prior^POS^ individuals (**FIG 5B**) and reacted against the Spike RBD domain (**FIG 5C**). These results indicate that although IgG^N^ and IgG^Spike^ both persist in saliva following natural infection, IgG^Spike^ persists at higher levels and reacts against Spike regions that are essential for ACE2 binding (i.e., the RBD).

### VII. Individuals with breakthrough CoV^Delta^ infections had comparable saliva IgG^Spike^ levels to those of uninfected, vaccinated individuals

During the period of Dec 2020 – Mar 2021, COVID vaccination was prioritized and available to the elderly and other individuals at increased risk of severe disease (e.g. healthcare workers, first responders). In Ohio, beginning on Mar 22 2021, individuals who were 16 years or older could receive a COVID vaccine, including all college students (*69*). Despite the widespread availability of vaccines by our Autumn 2021 semester, CoV2^Delta^ lineage infections occurred among unvaccinated (Vax^NEG^) individuals and vaccinated (Vax^POS^) individuals. The term “breakthrough infection” is older than COVID (*70*) but is now commonly applied to individuals who are PCR^POS^ despite their being Vax^POS^. Since BNT162b2, mRNA-1273 and Ad26.COV2.S were each designed to elicit an Ig response against CoV2 Spike (since it is essential for CoV2 infection of ACE2-expressing cells), we assessed whether breakthrough infections with CoV2^Delta^ were associated with lower levels of Spike-specific Ig in saliva compared to PCR (neg) vaccinates. Shown in **FIG 6** are saliva levels of IgM^Spike^, IgA^Spike^ and IgG^Spike^ in three groups of individuals: VAX^NEG^PCR^POS^ individuals infected with CoV2^Delta^, VAX^POS^PCR^POS^ individuals infected with CoV2^Delta^, and VAX^POS^PCR^NEG^ individuals. Saliva from VAX^NEG^PCR^POS^ and VAX^POS^PCR^POS^ individuals was collected during Wave 4 (**FIG 2**), when community viral burdens were their highest (**FIG 3D**); saliva from VAX^POS^PCR^NEG^ individuals was collected shortly after Wave 4 had passed. These results demonstrate that VAX^POS^PCR^POS^ and VAX^POS^PCR^NEG^ groups each had significantly higher saliva IgG^Spike^ levels than VAX^NEG^PCR^POS^ individuals (**FIG 6C**). Furthermore, the saliva IgG^Spike^ levels of VAX^POS^PCR^POS^ and VAX^POS^PCR^NEG^ groups did not significantly differ from one another (**FIG 6C**). Notably, although saliva IgM^Spike^ levels were indistinguishable across groups (**FIG 6A**), VAX^NEG^PCR^POS^ individuals were distinguished by the highest levels of saliva IgA^Spike^ (**FIG 6B**). Similar trends were observed using recombinant CoV2^Alpha^, CoV2^Beta^, CoV2^Gamma^ and CoV2^Delta^ Spike as capture antigens (**Supplemental FIG S6**). We conclude from this that COVID vaccination increased saliva IgG^Spike^ levels in our university community as intended, the saliva IgG^Spike^ levels in all vaccinees being comparable (regardless of whether they had a breakthrough CoV^Delta^ infection) and significantly higher than the saliva IgG^Spike^ levels of unvaccinated, infected individuals.

**FIGURE 6.**
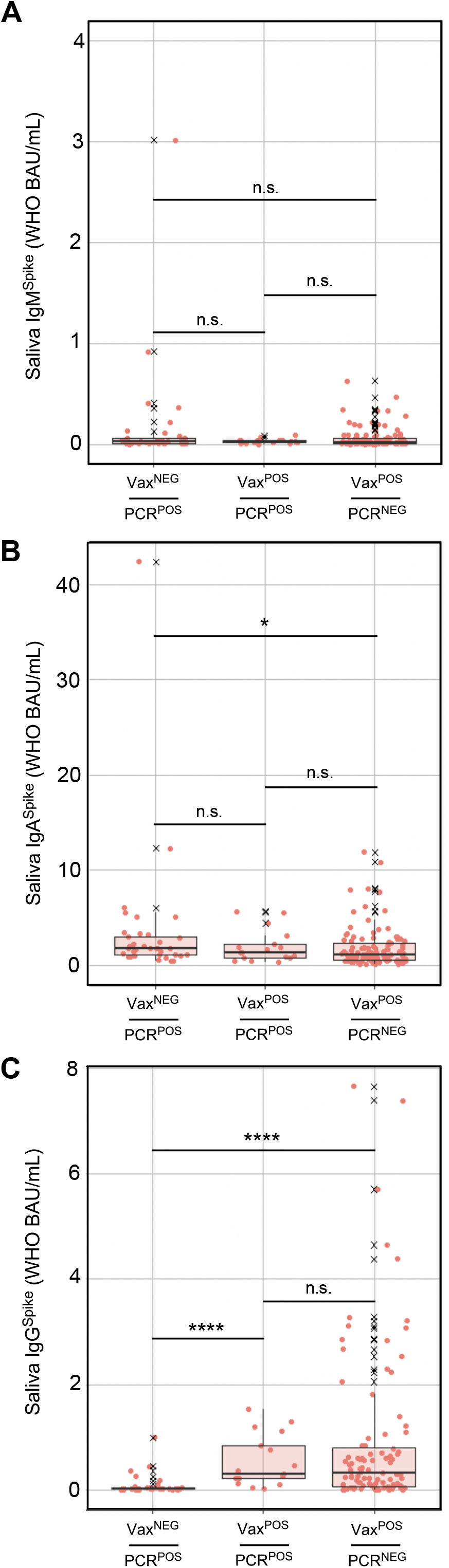
Spike-specific Ig levels in saliva of CoV2^Delta^-infected unvaccinated individuals, CoV2^Delta^-infected vaccinees, and uninfected vaccinees. During and shortly after COVID Wave 4 (i.e. that which was caused by CoV2^Delta^), saliva from three groups of individuals were collected and used for Ig measurements: those who had not been fully vaccinated and were positive for the CoV2^Delta^ lineage (Vax^NEG^PCR^POS^), those who had been fully vaccinated and were positive for the CoV2^Delta^ lineage (Vax^POS^PCR^POS^), and those who had been fully vaccinated and were negative for any CoV2 lineage (Vax^POS^PCR^NEG^). Shown are the (**A**) IgM^Spike^, (**B**) IgA^Spike^ and (**C**) IgG^Spike^ levels in each individual sample per group. ^X^, values that were considered outliers but are nevertheless shown for completeness and are included in all statistical group comparisons. * p≤ 0.05, as determined by unpaired Wilcoxon tests with Benjamini-Hochberg adjustment.

### VIII. Despite comparable Spike-specific Ig levels, CoV2^Delta^-infected vaccinee saliva was less capable of Spike:ACE2 inhibition, relative to uninfected vaccinees

Since the presence of CoV2-specific Ig does not equate to its having neutralization capacity (*71*), we next compared the ability of Vax^NEG^PCR^POS^, Vax^POS^PCR^POS^ and Vax^POS^PCR^NEG^ saliva samples to inhibit Spike:ACE2 interactions. We quantified inhibitory activity using an ACE2 displacement assay (**FIG 7A**), wherein plate-bound Spike was incubated with the same saliva samples above (i.e., those of **FIG 6**), followed by washing and addition of a luminescent probe-conjugated, recombinant form of human ACE2. The extent to which luminescence declined relative to non-saliva treated wells was used to derive a percent inhibition value for each individual sample (see *Methods* for additional details). The results of this analysis are shown in **FIG 7B** and demonstrate that there were differences between cohorts, the inhibitory activity of VAX^POS^PCR^NEG^ saliva being significantly higher than that of Vax^NEG^PCR^POS^ saliva (**FIG 7B**). The inhibitory activity of Vax^POS^PCR^POS^ saliva (median=12) was 50% higher than that of Vax^NEG^PCR^POS^ saliva (median=8), but 25% lower than that of Vax^POS^PCR^NEG^ saliva (median=16); as a whole, however, the inhibitory activity of Vax^POS^PCR^POS^ saliva did not significantly differ from that of Vax^NEG^PCR^POS^ saliva, nor did it significantly differ from Vax^POS^PCR^NEG^ saliva (**FIG 7B**). Within the Vax^NEG^PCR^POS^ cohort, there were no significant correlations between these samples’ inhibitory activity and their IgM^Spike^ (**FIG 7C**), IgA^Spike^ (**FIG 7D**) or IgG^Spike^ concentrations (**FIG 7E**). This was also true of the Vax^POS^PCR^POS^ cohort, as no significant correlations were observed between these samples’ inhibitory activity and their IgM^Spike^ (**FIG 7F**), IgA^Spike^ (**FIG 7G**) or IgG^Spike^ concentrations (**FIG 7H**). Within the Vax^POS^PCR^NEG^ cohort, there were significant correlations between samples’ inhibitory activity and their IgA^Spike^ concentration (**FIG 7J**), as well as their IgG^Spike^ concentration (**FIG 7K**), but not their IgM^Spike^ concentration (**FIG 7I**). When considered alongside the data shown in **FIG 6**, we conclude COVID vaccination led to increases in saliva IgG^Spike^ concentrations, the levels being similar between vaccinees who had a breakthrough CoV2^Delta^ infection (Vax^POS^PCR^POS^) and vaccinees who did not (Vax^POS^PCR^NEG^), but that during Wave 4 the antibodies in Vax^POS^PCR^POS^ saliva were limited in their ability to inhibit Spike, the inhibition values being intermediate between Vax^POS^PCR^NEG^ saliva (which had the highest inhibition values) and Vax^NEG^PCR^POS^ controls (which had the lowest inhibition values).

**FIGURE 7.**
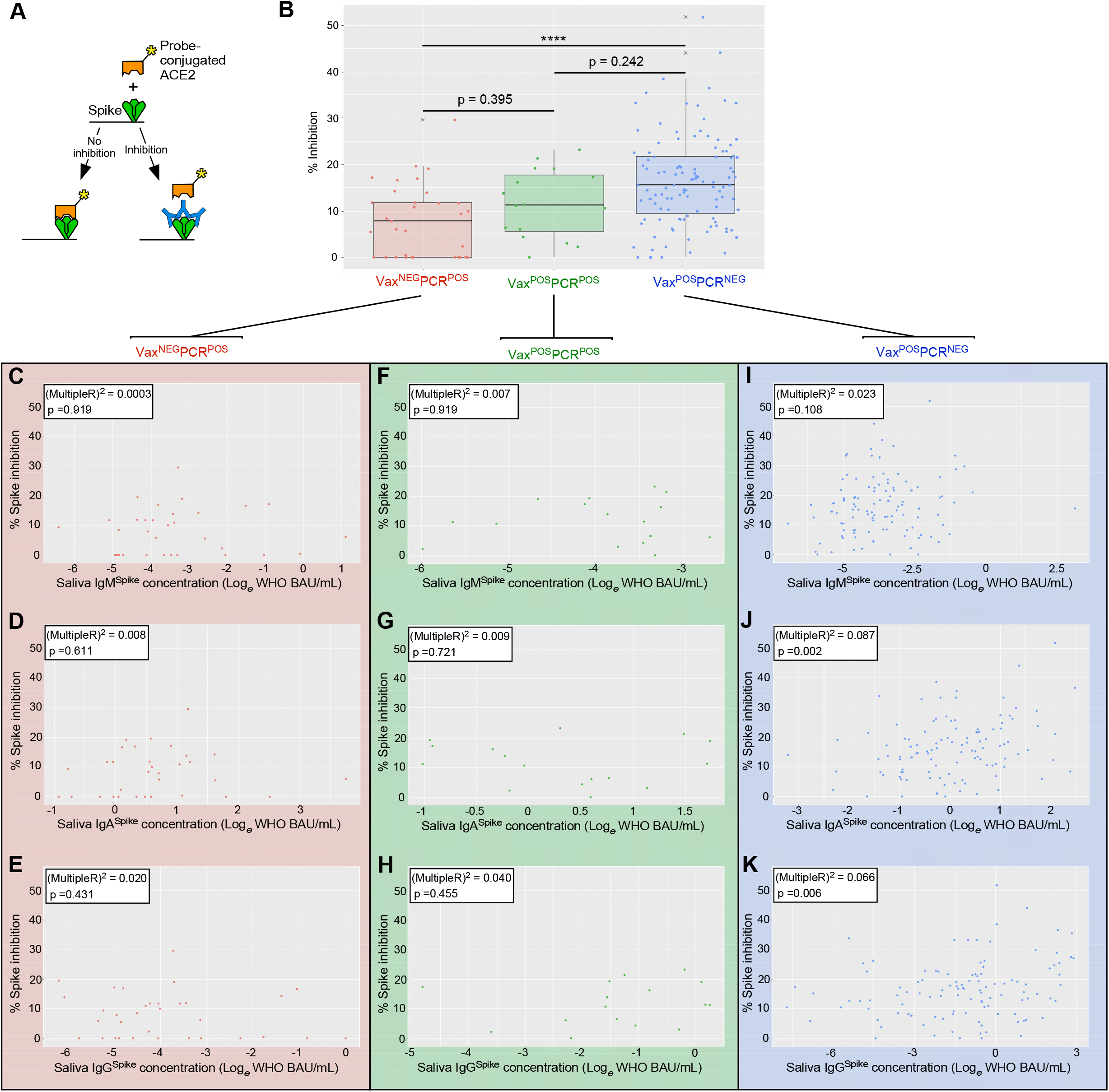
Inhibition of Spike function by saliva of CoV2^Delta^-infected unvaccinated individuals, CoV2^Delta^-infected vaccinees, and uninfected vaccinees. **(A)** Depiction of the probe-conjugated ACE2 displacement assay used to measure saliva samples’ ability to inhibit CoV2 Spike binding to its human receptor, ACE2. The samples in this case were from VAX^NEG^PCR^POS^, VAX^POS^PCR^POS^ and VAX^POS^PCR^NEG^ individuals (the same samples used for IgM^Spike^, IgA^Spike^ and IgG^Spike^ measurements in **FIG 6** above). (**B**) The percent inhibition value of each individual sample in each group. Within the (**C-E**) VAX^NEG^PCR^POS^ group, (**F-H**) VAX^POS^PCR^POS^ group, and (**I-K**) VAX^POS^PCR^NEG^ group, the relationship between an individual samples’ inhibition value and cognate (**C**,**F**,**H**) IgM^Spike^ concentration, (**D**,**G**,**I**) IgM^Spike^ concentration, and (**E**,**H**,**J**) IgG^Spike^ concentration. Graph insets indicates the Multiple R-squared value associated with the linear regression model of the respective data set (i.e. the % variation in inhibition that can be explained by the indicated Ig concentration), as well as its p-value (i.e. the significance of the linear model as a whole).

## DISCUSSION

The spread of CoV2 to the US marked the beginning of an extraordinary period wherein a novel respiratory virus transmitted and evolved in a population with no prior immunity, our primary defenses being behavioral changes (e.g., masking and physical distancing) until the advent of effective vaccines. The first COVID case in the US occurred in January 2020 (*72*). It was soon discovered that CoV2 caused both symptomatic and asymptomatic infections (the latter being more common in young adults), that asymptomatic individuals could transmit CoV2 (*73, 74*), and that isolation of symptomatic individuals alone would not sufficiently “flatten the curve” of COVID incidence (*37, 75*). By April 2020, most US universities shut down on-campus activities so as to limit CoV2 transmission among their students, staff, and faculty. Many universities established COVID monitoring programs prior to campus reopening as a means of identifying symptomatic and asymptomatic individuals. These monitoring programs varied in their testing modalities (PCR-or antigen-based), cadence (weekly versus biweekly testing) and sample pooling practices (pooled versus individual testing); all monitoring programs, however, had the same goal in mind: enabling safe resumption of on-campus classes and activities. Now that mass COVID testing programs have ended in US, enabling time for processing and reflection, we are sharing the results of our monitoring program which we believe are most relevant to the ongoing issues of community spread, the longevity of mucosal Ig following natural infection, breakthrough infections, and the utility of saliva for assessing Ig responses to newer Omicron subvariants and booster vaccines.

That the COVID waves in our campus community mirrored those which occurred in surrounding counties, instead of preceding the surrounding county waves, touches on an important question at the time regarding campus reopening: what if any contribution would the influx of students have on COVID incidence in surrounding communities. In January 2021, student returns to university campuses were a contentious subject in the US due to the potential risk of contracting the virus and subsequent transmission to surrounding communities. COVID vaccines were not yet widely available to young adults, and—fairly or unfairly—university students were perceived as being more cavalier in their adherence to masking protocols and social distancing. Whether or not the reopening of a given college or university contributed to higher off-campus COVID transmission will depend on several variables (e.g. whether a school was in a state that mandated mask-wearing) (*76*), but in our case the COVID wave that occurred in our university in January 2021 (Wave 1) peaked during the tail end of one which had been ongoing in surrounding counties (compare **FIG 2** to supplemental **FIG S2**). This was also true in Aug 2022, when our campus reopened after summer break and experienced Wave 4, which followed the Delta wave that had already begun in surrounding counties. The timing of Wave 1 and Wave 4 in relation to those in surrounding counties is inconsistent with the argument that our university reopening contributed to COVID incidence in the surrounding communities. Studies at other large universities with COVID policies and monitoring programs similar to our own support this conclusion (*77-79*).

Early in the COVID pandemic, it was unknown whether natural infection would give rise to Ig responses that were durable and protective, as those against common seasonal coronaviruses are short-lived (only 6 months in some cases) (*80*), or worse still whether the Ig response would actually enhance infection or disease (*81-84*). Regarding the durability and protective capacity of the antibody response to natural CoV2 infection, current knowledge on this subject was recently reviewed (*71*). In our study, at the time of initial PCR positivity we could already detect elevations in CoV2-specific Ig (IgM, IgA, and to a lesser extent IgG) in the saliva of asymptomatic individuals, the degree to which varied by lineage, CoV2^Delta^ being the most immunogenic of the lineages we assessed. Saliva levels of CoV2-specific IgG were substantially higher in Prior^POS^ individuals compared to New^POS^ individuals, were directed against Spike, Spike RBD and (to a lesser extent) the N protein. Potential reasons why Spike-specific IgG (IgG^Spike^) levels were higher than those of N-specific IgG (IgG^N^) include Spike being more antigenic, or alternatively it may reflect an inherent inability of IgG^N^ to persist in saliva relative to IgG^Spike^, as is the case in plasma (*85*). With regards to antibody dependent enhancement (ADE) of virus infection or subsequent disease, this has primarily been studied in the context of flaviviruses (*86*) and is the phenomenon wherein non-neutralizing antibodies amplify viral entry (instead of blocking it). ADE occurs in cell culture and animal infection models of SARS-CoV-1 (*87, 88*) and MERS-CoV (*89*), as well as feline infectious peritonitis coronavirus (FIPV) (*90, 91*). B cells from convalescent COVID patients can produce monoclonal antibodies that enhance the ACE2-binding capacity of Spike and CoV2 infectivity in cell culture (*92, 93*); however, these same monoclonal antibodies did not enhance CoV2 infection in mouse or macaque models (*92*), nor has evidence of vaccine-enhanced disease (VED) been observed in the hamster, ferret or macaque COVID models (*94, 95*). The circumstances and extent to which ADE occurring following CoV2 infection nevertheless remains an active area of research (*96-98*).

When COVID vaccine doses were in short supply (early 2021), university students were generally not considered a vaccine priority by national public health agencies. By the time COVID vaccines were widely available, non-trivial levels of vaccine hesitancy had arisen among university students in many countries for many reasons (*99*). Vaccine hesitancy was reinforced by the occurrence of breakthrough infections with CoV2^Delta^ (*100, 101*), the first lineage to emerge after vaccines had become more widely available in Summer 2021. If vaccines were effective, conventional logic at the time being, how then could a vaccinated individual still become PCR^POS^? Our current understanding is that a combination of three factors affects susceptibility to breakthrough infections: (1) antibody levels at the time of virus exposure, (2) the neutralizing capacity of these antibodies, and (3) the amount of virus to which a vaccinee is exposed. Our data demonstrate that saliva IgG^Spike^ levels were comparable between CoV^Delta^-infected vaccinees (Vax^POS^PCR^POS^) and uninfected vaccinees (Vax^POS^PCR^NEG^), but that the collective inhibitory capacity of this IgG^Spike^ and other saliva antibodies differed between groups, with Vax^POS^PCR^POS^ saliva being less inhibitory than Vax^POS^PCR^NEG^ saliva (**FIG 7B**). If the saliva Ig response is representative of that which occurs in other parts of the upper airway, then the combination of weak neutralization capacity and higher viral loads, which were typical of the Delta wave (Wave 4 of **FIG 3D**), created conditions that were conducive to CoV2^Delta^ breakthrough infections. Our observation that CoV2^Delta^ was more concentrated in saliva of asymptomatic individuals is consistent with work showing CoV2^Delta^-infected individuals were more likely to transmit virus before developing symptoms, compared to individuals infected with pre-Delta lineages (*102*).

The largest COVID wave our university community experienced was caused by the Omicron lineage. The Omicron lineage spread rapidly after its first detection in southern Africa in November 2021 (*11, 103*); the >30 amino acid substitutions in Spike enabled Omicron to bind ACE2 with higher affinity, as well as escape the anti-Spike antibody response elicited by either natural infection or vaccination with pre-Omicron lineages or vaccines (*104-106*). The immunoevasive properties of Omicron are consistent with its causing a COVID wave in our community after vaccine mandates had been established. The rapidity with which Omicron took over was observed in other university settings which, like ours, were highly vaccinated at the time (*107*). Compared to infections caused by the Delta lineage, those by Omicron tend to cause less severe disease (*108*), which may be due in whole or part to its being enriched in upper airways (including the oral cavity) as opposed to the lower airways (*39-43*). Omicron variants BA.1 and BA.2 were the last lineages detected in our university community before our testing program ended in May 2022. Since then CoV2 has continued to evolve, there now being additional Omicron variants (BA.4, BA.5, BA.2.12.1, BA.2.75, XBB) and “Scrabble” subvariants (BQ.1 and BQ1.1) with Spike protein sequences that further desensitize the virus to *in vitro* neutralization by many (but not all) monoclonal therapies (*109-112*), as well as convalescent plasma (*113*). Since Omicron has a higher tropism for the nasopharyngeal and oral cavities than that of pre-Omicron lineages (*39-43*), saliva antibodies may be more important inhibitors of Omicron transmission than plasma or lower airway antibodies, and saliva—the collection of which is far easier than blood—may be more suitable for rapid determination of whether someone has neutralizing capacity against future CoV2 variants that have yet to emerge.

The limitations of our study are as follows: (1) Since participants in our monitoring program provided saliva on a weekly basis, we cannot know the exact date on which someone was infected, rather only that they were infected 0-7 days prior to their scheduled test; (2) By only measuring CoV2-specific Ig in individuals whose C_T_ values fell within a narrow range (thus normalizing for viral load), we cannot make any statements regarding the relationship between lower or higher C_T_ values and CoV2-specific Ig levels; (3) Although we can correlate saliva samples’ Spike inhibition capacity with their corresponding IgM^Spike^, IgA^Spike^ and IgG^Spike^ levels, we cannot definitively state which of these isotypes most contributed to inhibition; (4) Finally, we did not measure CoV2-specific Ig levels in individuals infected with CoV2^O-BA.1^ or CoV2^O-BA.2^, a reason being at that stage in the pandemic (i.e. Waves 5-6 in our community) vaccine mandates were in place, and boosters were becoming available, making it difficult if not impossible to discern what levels of IgM^Spike^, IgA^Spike^ and IgG^Spike^ were due to vaccination versus boosters versus Omicron infection.

## ACKNOWLEDGEMENTS

We would like to acknowledge the hundreds of Ohio State staff and student employees who collected, transported and processed saliva during the period that our campus testing program was active. We would also like to acknowledge the research compliance expertise of Tish Denlinger (Office of Responsible Research Practices) and Angela Emerson (Office of Research), the technical expertise of Dr. Arpita Agrawal, Brian Gribble, and Trina Wemlinger (Center for Clinical and Translational Science), the general lab assistance of Sam Hagenbaugh and Elizabeth Griffin (TOPS Program), and documentation assistance of Marie Klever (Infectious Disease Institute). This work was supported by the Analytical and Development Laboratory of the Clinical Research Center/Center for Clinical Research Management of Wexner Medical Center, the OSU College of Medicine (Microbial Infection & Immunity Department), OSU College of Public Health (Epidemiology Division), and the OSU Infectious Disease Institute.

---

**Supplemental FIGURE S1.**
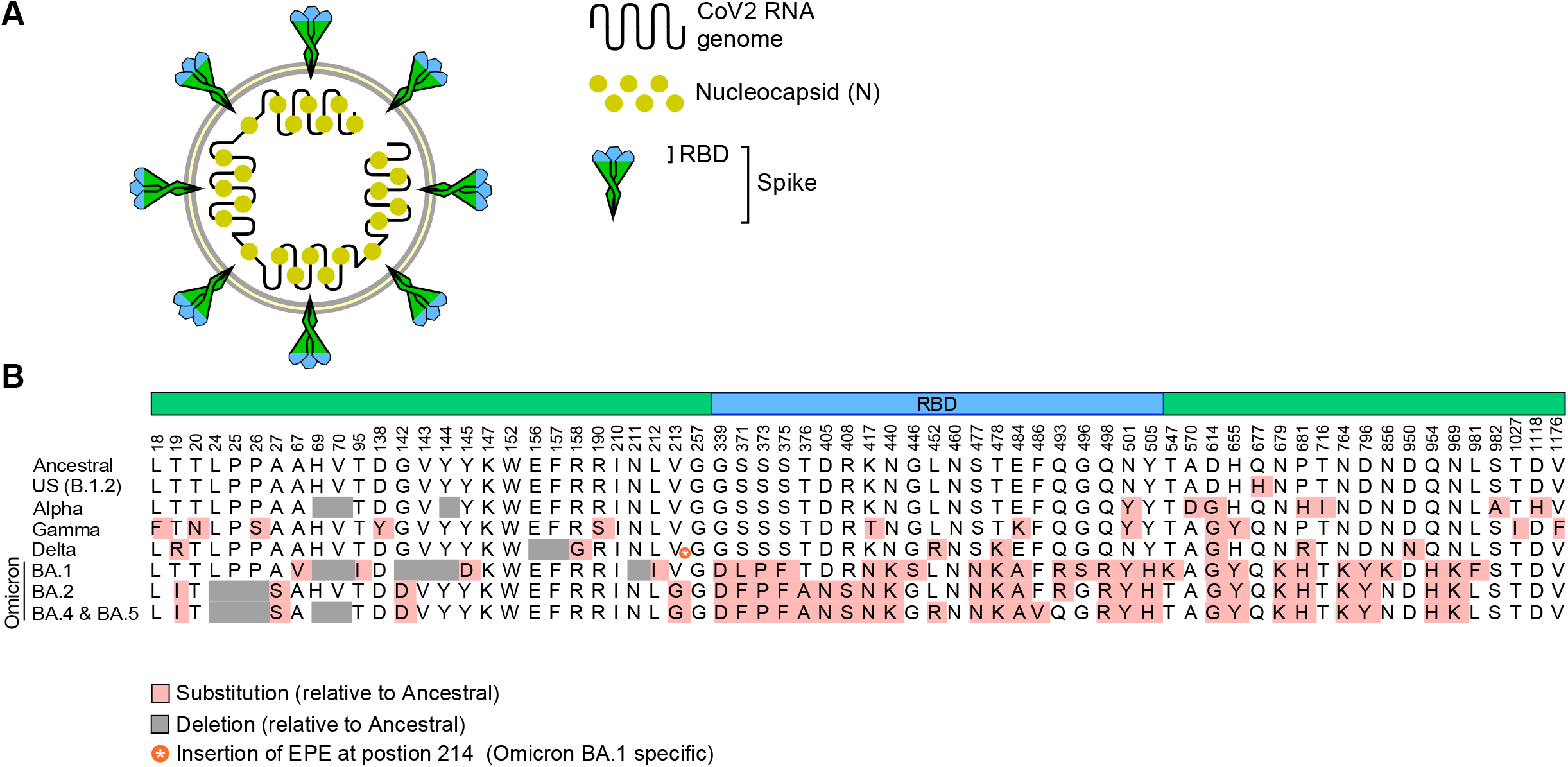
The CoV2 antigens and components relevant to our study. (**A**) Depiction of CoV2 and its RNA genome, nucleocapsid (N, yellow) and Spike proteins, the latter being differentially colored to indicate the Receptor Binding Domain (RBD, blue) and non-RBD regions (green). (**B**) The amino acids which distinguish the CoV2^Anc^ Spike protein from CoV2^US^ (also known as B.1.2), CoV2^Alpha^, CoV2^Gamma^ and CoV2^Delta^, as well as the Omicron lineages CoV2^O-BA.1^, CoV2^O-BA.2^, CoV2^O-BA.4^ and CoV2^O-BA.5^.

**Supplemental FIGURE S2.**
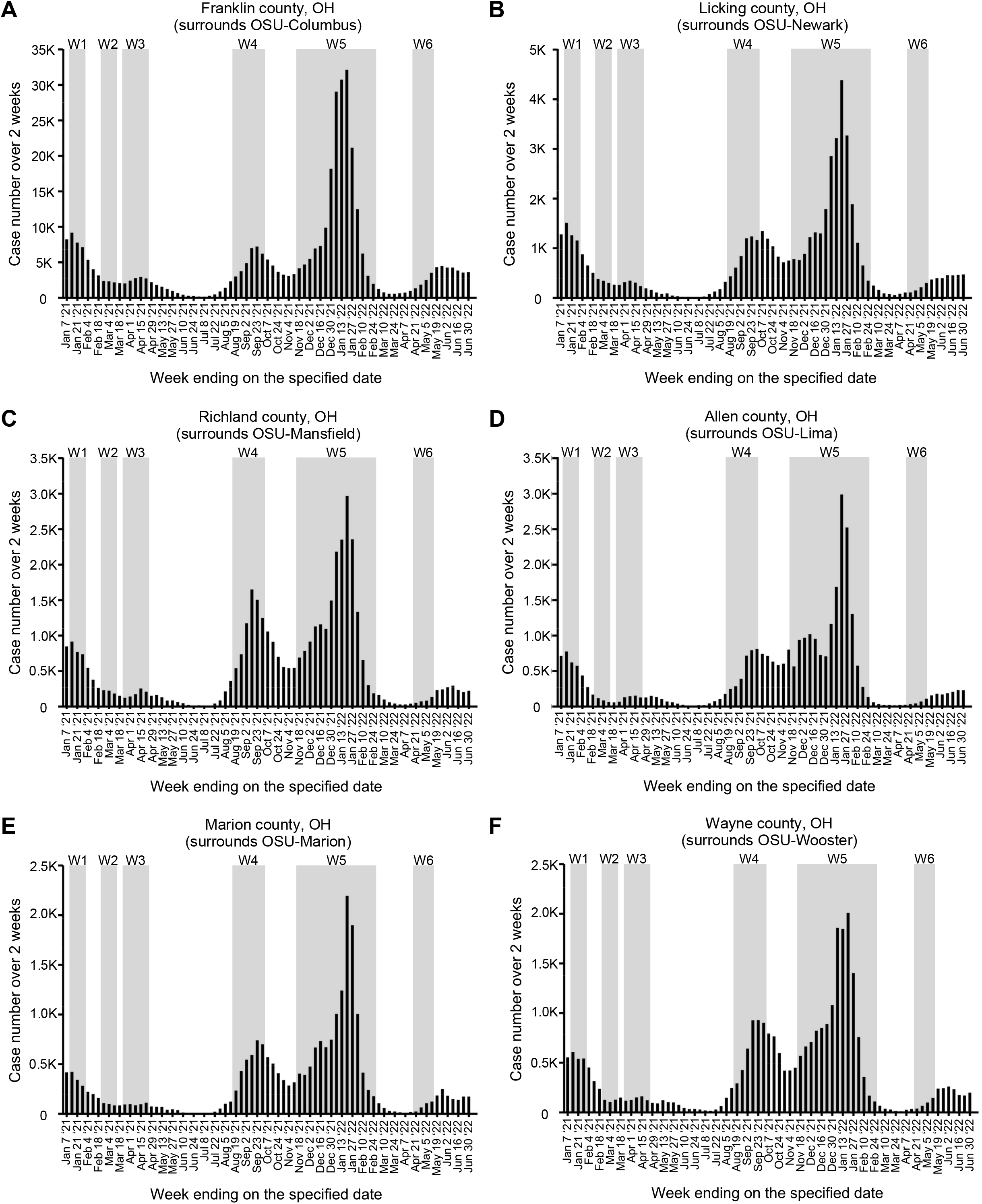
The waves of COVID incidence in the counties surrounding our university campuses. Daily COVID cases in the counties surrounding each campus of our university, as reported by the Ohio Department of Health (ODH), for the period spanning Jan 2021 → May 2022. Shown are the data for (**A**) Franklin County, which surrounds the OSU-Columbus campus; (**B**) Licking County, which surrounds the OSU-Newark campus; (**C**) Richland County, which surrounds the OSU-Mansfield campus; (**D**) Allen County, which surrounds the OSU-Lima campus; (**E**) Marion County, which surrounds the OSU-Marion campus; and (**F**) Wayne County, which surrounds the OSU-Wooster campus. Overlaid onto each graph are the dates which correspond to the six COVID waves (W1-W6) that occurred in our campus community (see **FIG 2**).

**Supplemental FIGURE S3.**
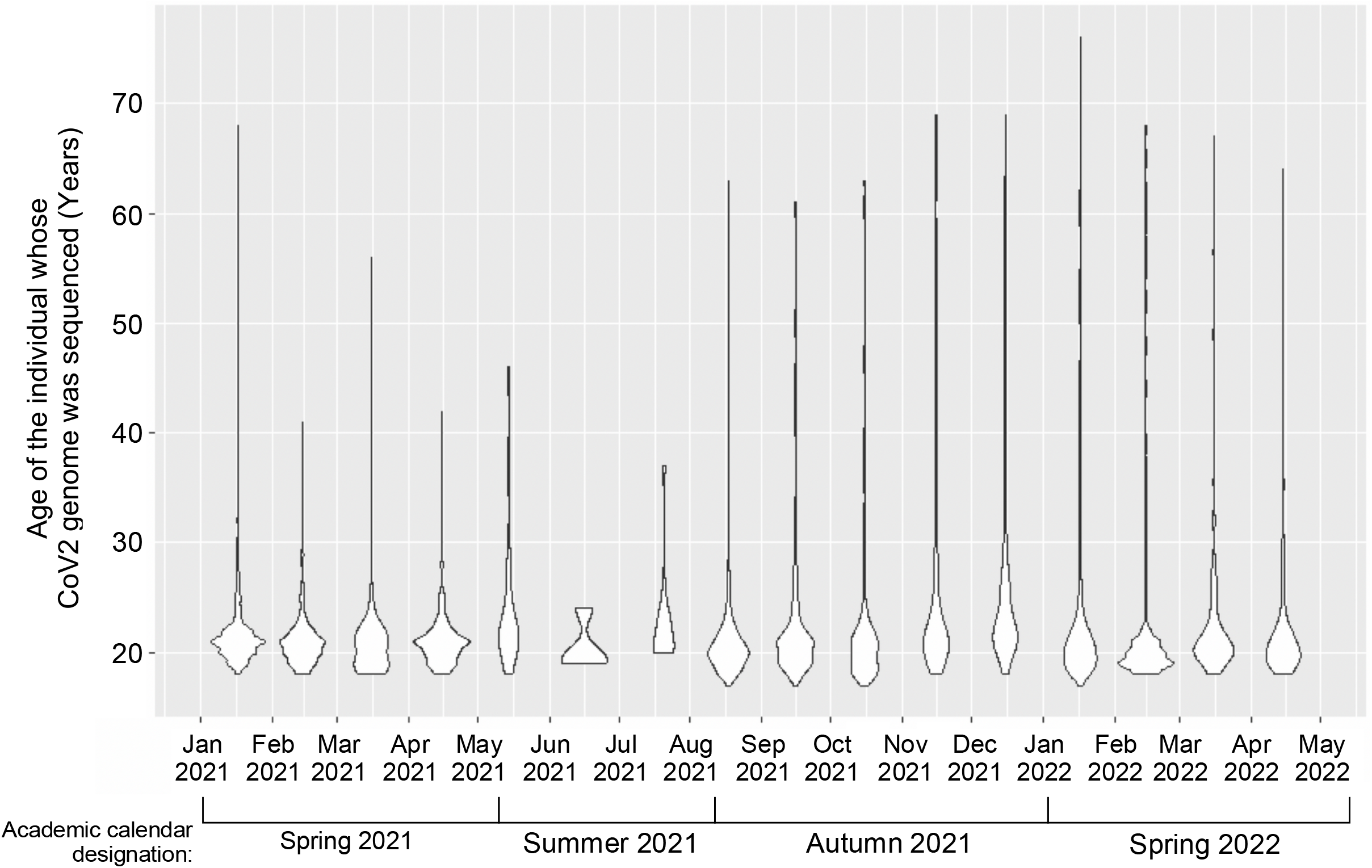
The age of individuals whose PCR^POS^ saliva sample was viral genome sequenced. The age range of individuals whose saliva was PCR^POS^ and sequenced throughout the monitoring period. Violin plots depicting the ages of individuals whose saliva was PCR^POS^ and sequenced each month.

**Supplemental FIGURE S4.**
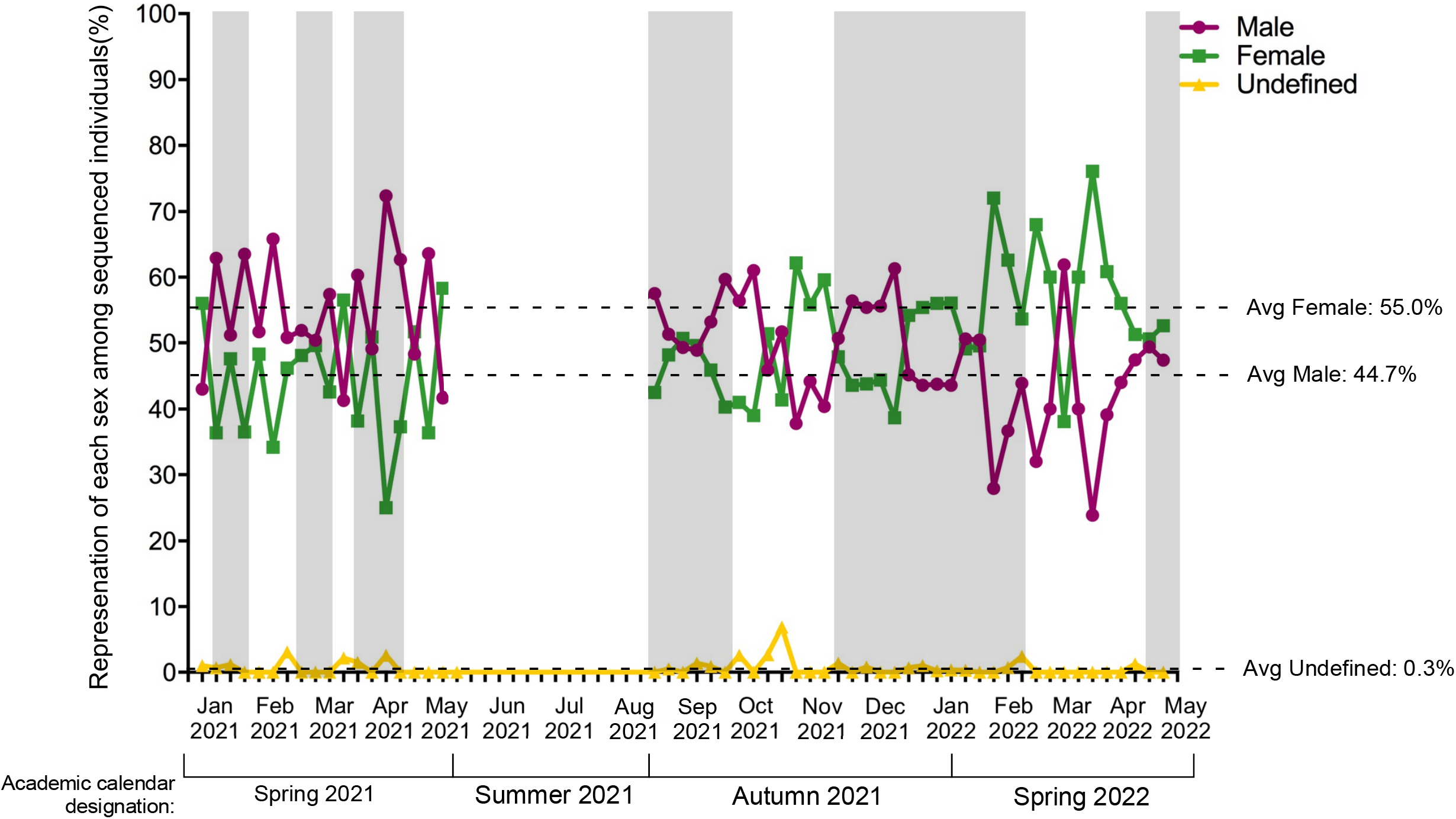
The representation of each sex among individuals whose PCR^POS^ saliva met sequencing criteria. The percent of males, females and undefined sex among individuals whose saliva was PCR^POS^ and sequenced for lineage identification for each week of our study period, the criteria for sequencing being a C_T_ ≤ 33. Overlaid onto the graph in gray are the periods corresponding to Waves 1-6 in our university community. The average values for each sex across the entire study period are indicated by the hatched lines.

**Supplemental FIGURE S5.**
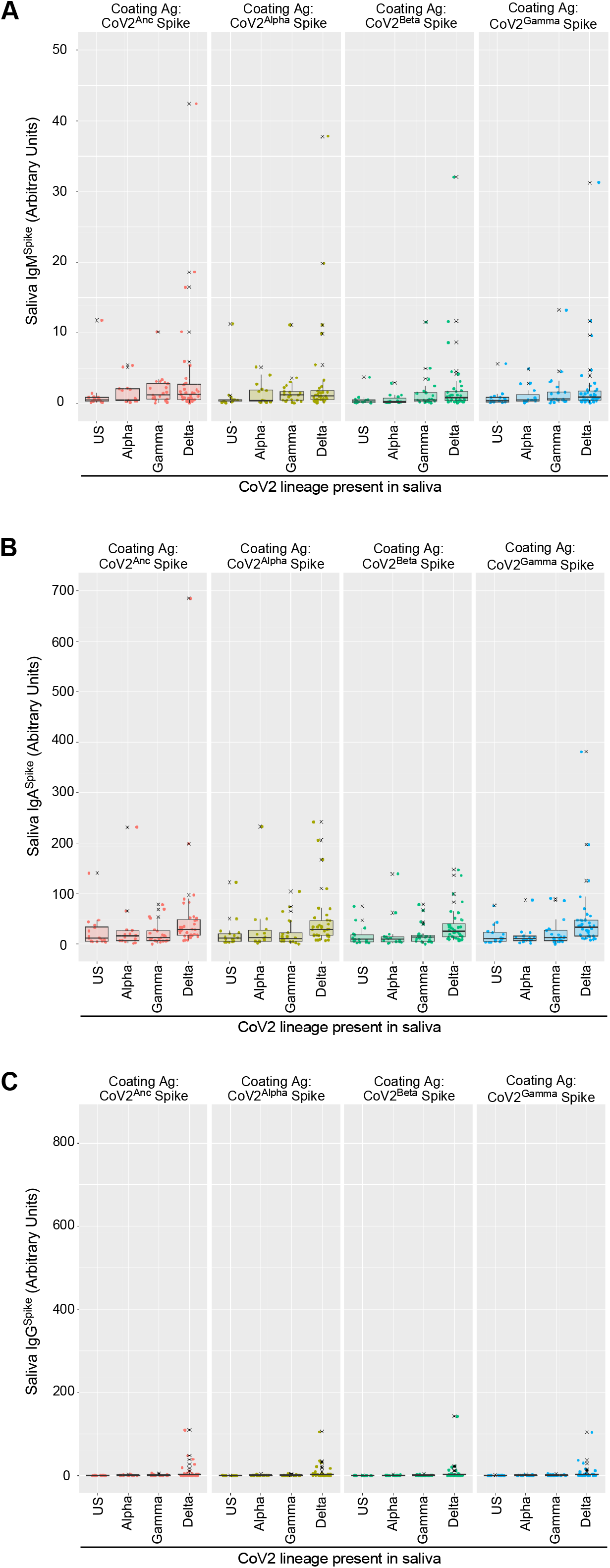
VOC Spike-specific Ig levels in the saliva of newly positive, asymptomatic individuals at the time of PCR testing. Saliva samples that were positive for either the CoV2^US^, CoV2^Alpha^, CoV2^Gamma^ or CoV2^Delta^ lineage were used to measure the concentrations of (**A**) IgM^Spike^, (**B**) IgA^Spike^ and (**C**) IgG^Spike^. Varying by column were the coating antigens (Ag) used for each measurement, the Ag being recombinant forms of either the CoV^Anc^ Spike (Column 1), CoV2^Alpha^ Spike (Column 2), CoV2^Beta^ Spike (Column 3), and CoV2^Gamma^ Spike (Column 4). Antibody levels are expressed in arbitrary units of luminescence. Note that the CoV^Anc^ -specific IgM, IgA and IgG values in (**A-C**) Column 1 were transformed into WHO Binding Antibody Units (BAUs) for **FIG 4**.

**Supplemental FIGURE S6.**
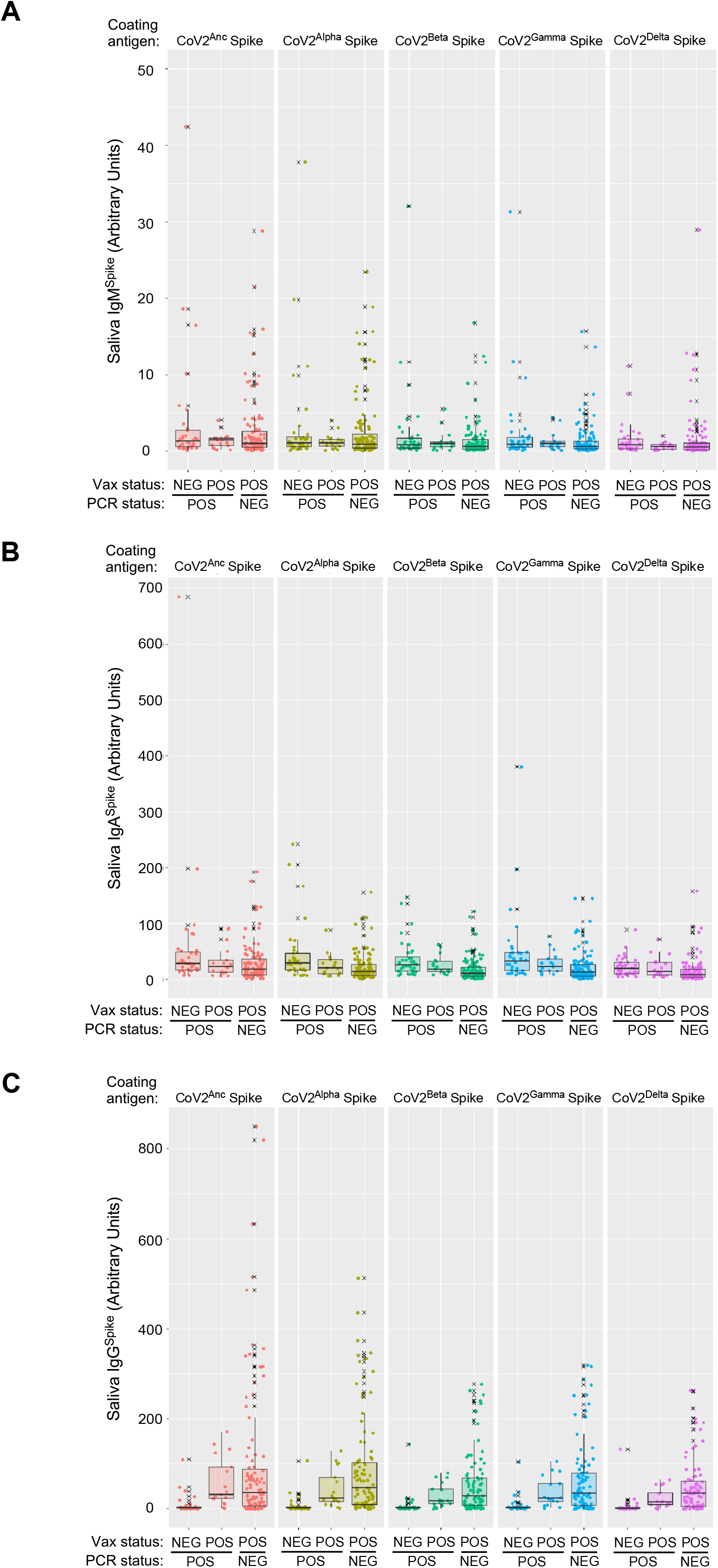
VOC Spike-specific Ig levels in saliva of CoV2^Delta^-infected unvaccinated individuals, CoV2^Delta^-infected vaccinees, and uninfected vaccinees. During and after COVID Wave 4 (i.e. that which was caused by CoV2^Delta^), saliva from three groups of individuals were collected and used for Ig measurements: those who had not been fully vaccinated and were positive for CoV2^Delta^ (Vax^NEG^ PCR^POS^), those who had been fully vaccinated and were positive for the CoV2^Delta^ (Vax^POS^ PCR^POS^), and those who had been fully vaccinated and were negative for any CoV2 lineage (Vax^POS^ PCR^NEG^). Shown for each individual in each group are the levels of (**A**) IgM^Spike^, (**B**) IgA^Spike^ and (**C**) IgG^Spike^ which bind to four different coating antigens (Ag), the Ag being recombinant forms of either the CoV^Anc^ Spike (Column 1), CoV2^Alpha^ Spike (Column 2), CoV2^Beta^ Spike (Column 3), CoV2^Gamma^ Spike (Column 4) and CoV2^Delta^ Spike (Column 5). Antibody levels are expressed in arbitrary units of luminescence. Note that the CoV^Anc^ -specific IgM, IgA and IgG values in (**A-C**) Column 1 were transformed into WHO Binding Antibody Units (BAUs) for **FIG 6**.

